# Reliable and generalizable brain-based predictions of cognitive functioning across common psychiatric illness

**DOI:** 10.1101/2022.12.08.22283232

**Authors:** Sidhant Chopra, Elvisha Dhamala, Connor Lawhead, Jocelyn A. Ricard, Edwina R. Orchard, Lijun An, Pansheng Chen, Naren Wulan, Poornima Kumar, Arielle Rubenstein, Julia Moses, Lia Chen, Priscila Levi, Alexander Holmes, Kevin Aquino, Alex Fornito, Ilan Harpaz-Rotem, Laura T. Germine, Justin T. Baker, BT Thomas Yeo, Avram J. Holmes

## Abstract

A primary aim of precision psychiatry is the establishment of predictive models linking individual differences in brain functioning with clinical symptoms. In particular, cognitive impairments are transdiagnostic, treatment resistant, and contribute to poor clinical outcomes. Recent work suggests thousands of participants may be necessary for the accurate and reliable prediction of cognition, calling into question the utility of most patient collection efforts. Here, using a transfer-learning framework, we train a model on functional imaging data from the UK Biobank (n=36,848) to predict cognitive functioning in three transdiagnostic patient samples (n=101-224). The model generalizes across datasets, and brain features driving predictions are consistent between populations, with decreased functional connectivity within transmodal cortex and increased connectivity between unimodal and transmodal regions reflecting a transdiagnostic predictor of cognition. This work establishes that predictive models derived in large population-level datasets can be exploited to boost the prediction of cognitive function across clinical collection efforts.

## Introduction

A key goal of precision psychiatry is the development of predictive models that provide personalized and robust estimates of clinically relevant phenotypes that can be used for prognostic and treatment decision making. To achieve clinical utility, these models must be accurate, and generalize across samples, diagnoses, measurement methods, and data-processing strategies. A primary barrier to progress in this area has been the historical use of small sample sizes, which has resulted in inflated prediction accuracies that largely fail to generalize across samples, populations, or collection sites^1–3^. Here, we demonstrate a modelling strategy that uses objective measurements of brain function to robustly predict global cognitive function across multiple transdiagnostic samples, yielding generalizable models between cohorts despite modest sample sizes. The models also simultaneously provide interpretable insight into the neurobiology of global cognitive functioning in common psychiatric illness.

Impaired cognitive functioning is a transdiagnostic characteristic of psychiatric illness^4–6^. It is difficult to treat^7,8^, strongly predicts social, occupational and functional impairment^9–11^, and is widely regarded by patients as a key priority for treatment^12,13^. Performance on cognitive tasks has repeatedly been linked to the structural and functional integrity of regions within transmodal association cortices. These regions are responsible for the integration of multiple sources of interoceptive and exteroceptive information and believed to underpin “higher-order” associative processes which support cognition untethered from immediate sensory inputs^14–16^, including adaptive goal-directed behavior^17^, the application of complex rules^18^, and the dynamic control of motor outputs^19^. Across patient populations, converging evidence suggests the presence of altered functioning within the large-scale systems that comprise association cortex^6,20–24^. In particular, impaired connectivity within the default network, encompassing aspects of medial prefrontal, posterior/retrosplenial, and inferior parietal cortices, has been observed across diagnostic categories^20,25–28^, while the level of dysconnectivity in the frontoparietal network, encompassing aspects of the dorsolateral prefrontal, dorsomedial prefrontal, lateral parietal, and posterior temporal cortices^29^ often tracks the severity of diagnoses and observed cognitive deficits^30–32^. However, despite the importance of establishing network-level predictors of symptom severity, the extent to which individual-specific profiles of brain functioning track clinically relevant cognitive impairments remains to be determined.

Inter-regional functional coupling of hemodynamic signals measured with functional Magnetic Resonance Imaging (fMRI), here termed functional connectivity, has recently emerged as a powerful and robust predictor of global cognitive functioning in healthy populations^33–37^. However, population neuroscience studies suggest that sample sizes exceeding thousands of participants may be required to develop accurate and stable brain-based predictive models of behavior^2,38–40^. This requirement far exceeds the vast majority of samples available to psychiatric research groups, calling into question both the utility and feasibility of developing clinically focused predictive models. Moreover, even brain-cognition predictive models derived from consortia-level samples can fail to generalize or show substantially reduced accuracy when applied to different datasets^2,38,41–43^, greatly diminishing the scope of their potential applications. In large population-based cohorts, the functioning of specific brain systems can be leveraged to predict a broad variety of phenotypes, ranging from demographic factors to physical and mental health-related variables^44–47^. The associated brain-based models, which are derived from tens of thousands of healthy individuals, likely contain information that could be translated to smaller clinical cohorts, allowing for the prediction of illness- and treatment-relevant phenotypes. In this regard, a recently developed framework called ‘meta-matching’^47^ capitalizes on the fact that a limited set of overlapping functional circuits are associated with a wide variety of phenotypes and uses high throughput population-based collection efforts to boost predictions of phenotypes in smaller cohorts. Using this framework, we have previously demonstrated a substantial increase in prediction accuracies for a board range of variables in population-based healthy samples^47^. However, the extent to which the meta-matching approach can improve prediction of clinically relevant behaviors in small independent patient samples, yield generalizable predictions, and generate neurobiological insight, all remain to be determined.

Here, we use the meta-matching framework to develop an accurate, generalizable, and interpretable transdiagnostic model of global cognitive functioning in across a diverse set of psychiatric illnesses. We find that across multiple distinct datasets, the meta-matching model results in prediction accuracies that are statistically significant, superior to conventional models, and comparable to those observed in much larger population-level studies. Moreover, the derived models are generalizable, meaning that they maintain performance across independent datasets. The functional networks that drive prediction across the datasets consistently implicate increased connectivity within transmodal association networks and decreased connectivity between transmodal and unimodal cortices as a fundamental, transdiagnostic predictor of global cognition.

## Results

### Accurate and generalizable prediction of global cognitive functioning across psychiatric populations

Our overall aim was to develop an accurate and generalizable brain-based model that can predict global cognitive functioning in patients with psychiatric illness. To this end, we applied the recently developed meta-matching framework, which capitalizes on the fact that a limited set of overlapping functional circuits are associated with a wide variety of health, cognitive and behavioral phenotypes^47^. First, we used resting-state fMRI data from 36,848 participants from the UK Biobank to derive functional connectivity estimates between 419 brain regions (^48,49^). Next, we used these connectivity values to train a single fully connected feed-forward Deep Neural Network (DNN) to predict 67 observed health, cognitive and behavioral phenotypes in the UK Biobank.

Using the meta-matching approach^47^, we then adapted this trained DNN (from the UK Biobank) to predict global cognitive function scores in three independent transdiagnostic clinical datasets: 1) the Human Connectome Project Early Psychosis (HCP-EP; n = 145), which includes individuals diagnosed with affective and non-affective psychosis; 2) the Transdiagnostic Connectomes Project (TCP; n = 101), which largely includes individuals diagnosed with mood and anxiety disorders; and 3) the Consortium for Neuropsychiatric Phenomics (CNP; n = 224), which is comprised of individuals diagnosed with schizophrenia, bipolar disorder or ADHD. All three samples also included a subset of healthy participants without psychiatric diagnoses. For a full demographic and diagnostic breakdown of the samples, see supplementary Table 1. Global cognitive function scores were derived for each clinical dataset using principal component analysis on a range of neuropsychological tests (see supplementary Table 2). Of note, the test batteries varied across datasets, allowing for the assessment of model robustness to study design and associated phenotype selection. For details about the meta-matching approach, please see Methods in the current manuscript.

Our first aim was to determine whether the meta-matching approach can make accurate and statistically significant predictions within clinical samples. For each dataset, we trained the meta-matching model using a nested cross-validation procedure, where each clinical sample was split into 100 unique training (70%) and test (30%) sets and the full meta-matching model was implemented for each train/test split. Accuracy was assessed as the mean Pearson correlation between the observed and predicted global cognition scores across the 100 test sets and statistical significance was assessed using a permutation testing procedure (see Methods for details). As shown in Fig1B, the meta-matching approach yields statistically significant predictions (all *ps* < 0.05) across all three datasets, with mean prediction accuracies comparable to those found using much larger healthy samples^50^. We find the same pattern of results when using the coefficient of determination to evaluate model accuracy (SFig1). Further, we establish that the meta-matching models systematically perform better than a standard prediction method, where a baseline comparison model was trained to predict cognition directly from the clinical sample functional connectivity values, with the difference between comparison and meta-matching models reaching statistical significance (all *ps* < 0.05).

**Fig 1.**
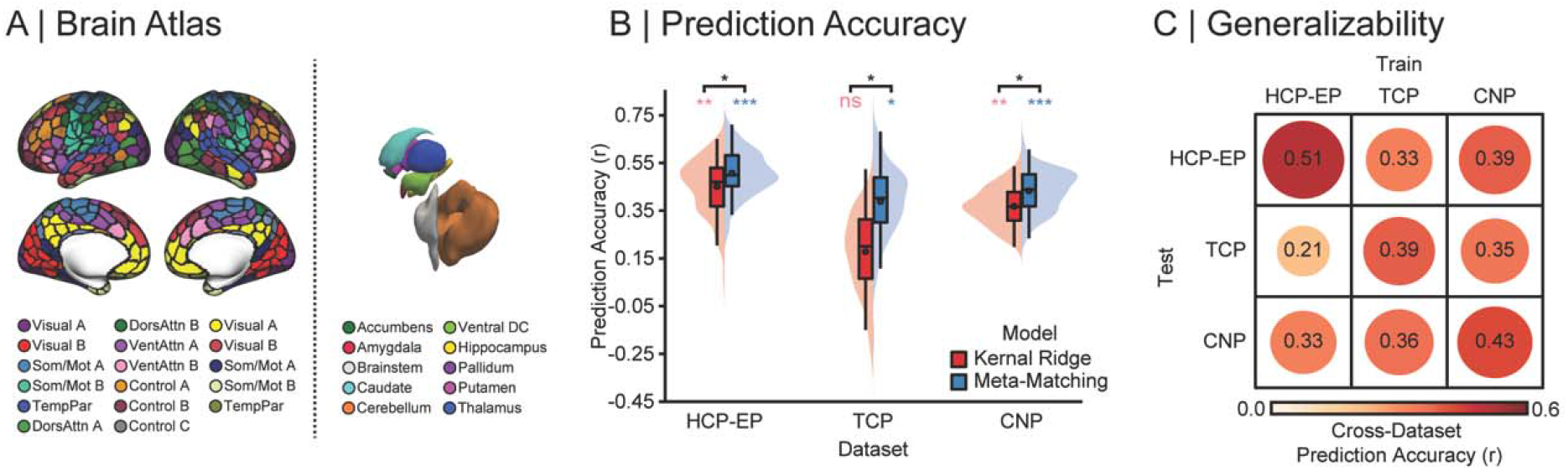
Accurate and generalizable prediction of global cognitive functioning across patient samples. **A)** The network organization of the human cortex. Colors reflect regions estimated to be within the same functional network according to the 17-network solution from Yeo, et al. ^51^ across the 400-parcel atlas from Schaefer, et al. ^48^, along with nineteen subcortical regions^49^. Cortical parcels and subcortical regions are used to extract BOLD timeseries and compute pair-wise functional connectivity estimates used for prediction. **B)** Prediction performance (Pearson’s correlation between observed and predicted values) using kernel ridge regression (red) and meta-matching (blue) across three transdiagnostic datasets: Human Connectome Project – Early Psychosis (HCP-EP), Transdiagnostic Connectomes Project (TCP) & UCLA Consortium for Neuropsychiatric Phenomics (CNP). Colored asterisks denote above-chance prediction (* = *p* < 0.05; ** = *p* < 0.001; *** = *p* < 0.0001; *ns = p* < 0.05) and black asterisks denotes statistically significant difference between models. **C)** Generalizability matrix for the meta-matching models, showing the prediction accuracy between the independent samples where the meta-matching model is trained in one dataset and then used to make predictions in an independent dataset. The diagonal represents the mean prediction performance within each dataset, which is also represented by the black dots in panel B.

Our second aim was to determine whether the meta-matching model generalizes across independent clinical collection efforts. Generalizability was assessed as the Pearson correlation between the observed and predicted global cognition scores when a model was trained in one dataset and tested on another dataset. Here, we trained the meta-matching model on the full sample of one dataset, and evaluated prediction accuracy on the other, resulting in six train-test prediction pairs between the three clinical datasets. Reflecting the presence of generalizable brain-behavior relationships across independent clinical cohorts, we observed that the meta-matching model generalizes across datasets (Fig1C) and reached prediction accuracies both comparable to the mean in-sample accuracies shown in Fig1B, and statistically significant for all but one train/test pair (train/test: HCP-EP/TCP). In all cases except this same train/test pair, higher generalizability was found when using the meta-matching model, compared to a standard prediction mode, with boosts in prediction accuracy ranging from 7% to 291% (SFig2). Scatterplots of observed and predicted values are provided in SFig3. We note that the meta-matching model generalized between datasets, where in addition to differences in diagnostic makeup, MRI scanner and acquisition parameters, cognition between each train-test pair were measured using different neurocognitive assessments, ranging from at-home online tests to gold-standard clinician-administered batteries.

### Predictive network features are stable across independent transdiagnostic datasets

We next determined the extent to which the neurobiological features that drive the predictions are shared between datasets. Predictive feature weights were derived using the Haufe transformation^52^. This transformation considers the covariance between functional connectivity and global cognition scores and, unlike regression coefficients, ensures that feature weights are not statistically independent of global cognition. It also increases both the interpretability and reliability of predictive features^52–54^. For each of the three datasets, we examined associations between average weights across the 100 cross-validation folds, at spatial scales of edges, regions, and networks. The edge-level spatial scale refers to the original 87,571 inter-regional pair-wise connections entered in the prediction models. By taking the mean of all edges attached to each of the 419 brain regions, edge-level connections can be aggregated into region-level predictive features. By taking the mean of all edges within and between 18 canonical functional networks including the subcortex (Fig1A; ^48^), edges-level connections can also be aggregated into 171 network-level predictive features. For both aggregated scales (region- and network-level), positive and negative feature weights were considered separately by zeroing negative or positive values before averaging, respectively.

When assessing associations between brain maps, spatial autocorrelation must be considered to ensure that any observed associations are not driven by low-level spatial properties of the brain^55^. This same consideration extends to associations between edge-level network maps, where connectively profiles of spatially adjacent regions demonstrate autocorrelation. To account for this property in the data we implemented a spin-test, which is a standardized procedure where the cortical regions of the atlas are rotated on an inflated sphere to generate configurations which preserve the spatial autocorrelation pattern of the cortex. We used these null atlas configurations to shuffle the rows and columns of the feature weight matrices to assess statistical significance of correlations between datasets.

Even after accounting for spatial autocorrelation, we find significant correlations across all three spatial scales (Fig2A-C). At the edge-level (Fig2A), we find low to moderate consistency between datasets, with the strongest correlation observed between the TCP and HCP-EP datasets (*r* = 0.31, *p*_*spin*_ < 0.001) and the TCP and CNP datasets (*r = 0*.*29, p*_*spin*_ < 0.001), with the CNP and HCP-EP datasets showing the weakest association (*r* = 0.14, *p*_*spin*_ < 0.001). At the region-level (Fig2B), we again find low to moderate consistency between datasets, with the strongest associations between datasets when examining negative feature weights (*rs* = 0.23 - 0.56; *ps*_*spin*_ < 0.001), indicating that regions where lower functional connectivity predicts better cognition are more highly related between datasets, relative to regions where higher functional connectivity predicted better cognition (*rs* = 0.01 - 0.24; *ps*_*spin*_ <0.001 - .998). The comparison between negative regional predictive features showing greater consistency between datasets than positive features was statistically significant for all three pairs of datasets (*Zs* = − 2.60 - 5.97, *ps*_*spin*_ < 0.009). At the network-level (Fig2C), we find the strongest overall consistency between datasets with moderate to high effect sizes observed when examining positive feature weights (*r* = 0.54 to 0.70; *ps*_*spin*_ < 0.001), indicating that network-level connections where lower functional connectivity predicts better cognition are strongly related between datasets, relative to network-level connections where higher functional connectivity predicted better cognition (*r* = 0.19 - 0.58; *ps*_*spin*_ < 0.001 - 0.250). The comparison between negative regional predictive features showing greater consistency between datasets than positive features was statistically significant for all three pairs of datasets (*Zs* = 2.88 - 5.75, *ps* < 0.004). Aggregating functional connectivity values at the canonical network-level capitalizes on the intrinsic functional architecture of the brain, with network-level brain function consistently being shown to have higher reliability^56,57^ compared to edge- and region-level measures. Therefore, aggregating features at the network-level may provide a more coherent signal than individual edge-level features, which may obscure associations between both individuals and datasets.

**Fig 2.**
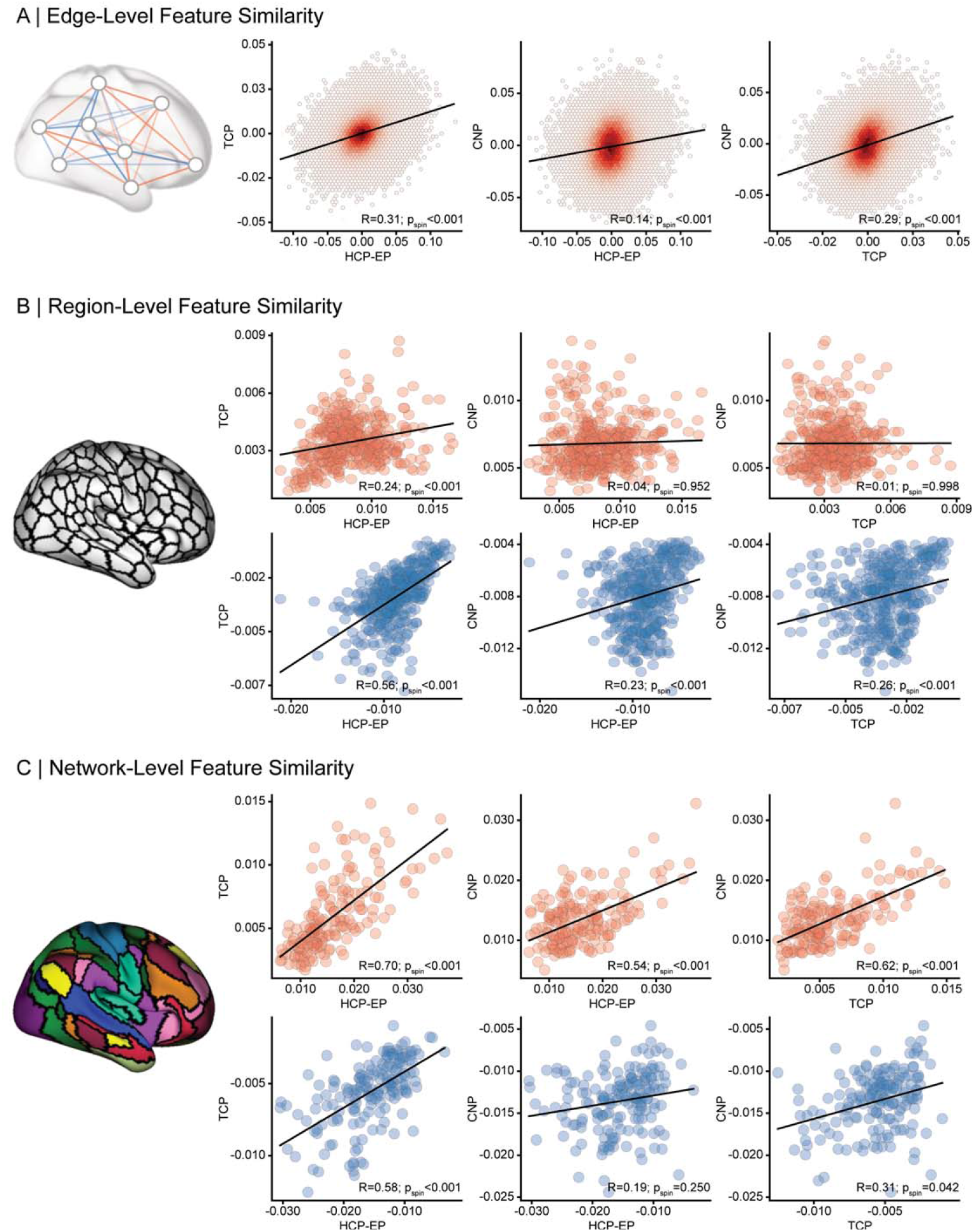
Predictive features are correlated between independent transdiagnostic datasets across scales. **A)** Association between Human Connectome Project – Early Psychosis (HCP-EP), Transdiagnostic Connectomes Project (TCP) & UCLA Consortium for Neuropsychiatric Phenomics (CNP) prediction model feature weights at the edge-level, which consist of 87,571 features per model. **B)** Association between feature weights of the three datasets at the region-level, where feature weights were averaged for all edges corresponding to a region, resulting in 419 regional features. Positive (red) and negative (blue) feature weights were considered separately by zeroing negative or positive values before averaging, respectively. All p-values displayed account for spatial autocorrelation between edges, regions, and networks. **C)** Association between feature weights of the three datasets at the network-level, where feature weights were averaged within and between each network, resulting in 171 network features per dataset. Positive and negative feature weights were considered separately by zeroing negative or positive values before averaging, respectively.

### Increased connectivity within transmodal systems and decreased connectivity between transmodal and unimodal systems predicts better cognitive functioning

Given that predictive features were most stable between datasets at the network-level, we examined the functional architecture of inter/intra-network connections driving prediction performance (Fig3A-C). In all three datasets, we observed a consistent, widespread, and complex pattern of network-level feature weights (Fig3B; *p*_FDR_ < 0.05). In line with prior work which reliably links functional coupling in transmodal association networks with cognition^24,58,59^, we find that brain-cognition relations converge on connections where higher functional connectivity within transmodal (default, frontoparietal and ventral attention) networks and lower functional connectivity between transmodal and unimodal (visual and somato/motor) networks predict better cognition (Fig3C). We also find that connectivity within the frontoparietal subnetwork A, encompassing aspects of dorsolateral prefrontal, lateral parietal, medial cingulate and posterior temporal cortices, was the strongest predictor of cognitive performance across datasets. More broadly, we find that in each of the three datasets, increased connectivity within unimodal, transmodal, as well as all aggregated cortical networks was predictive of better cognition (Fig 4).

**Fig 3.**
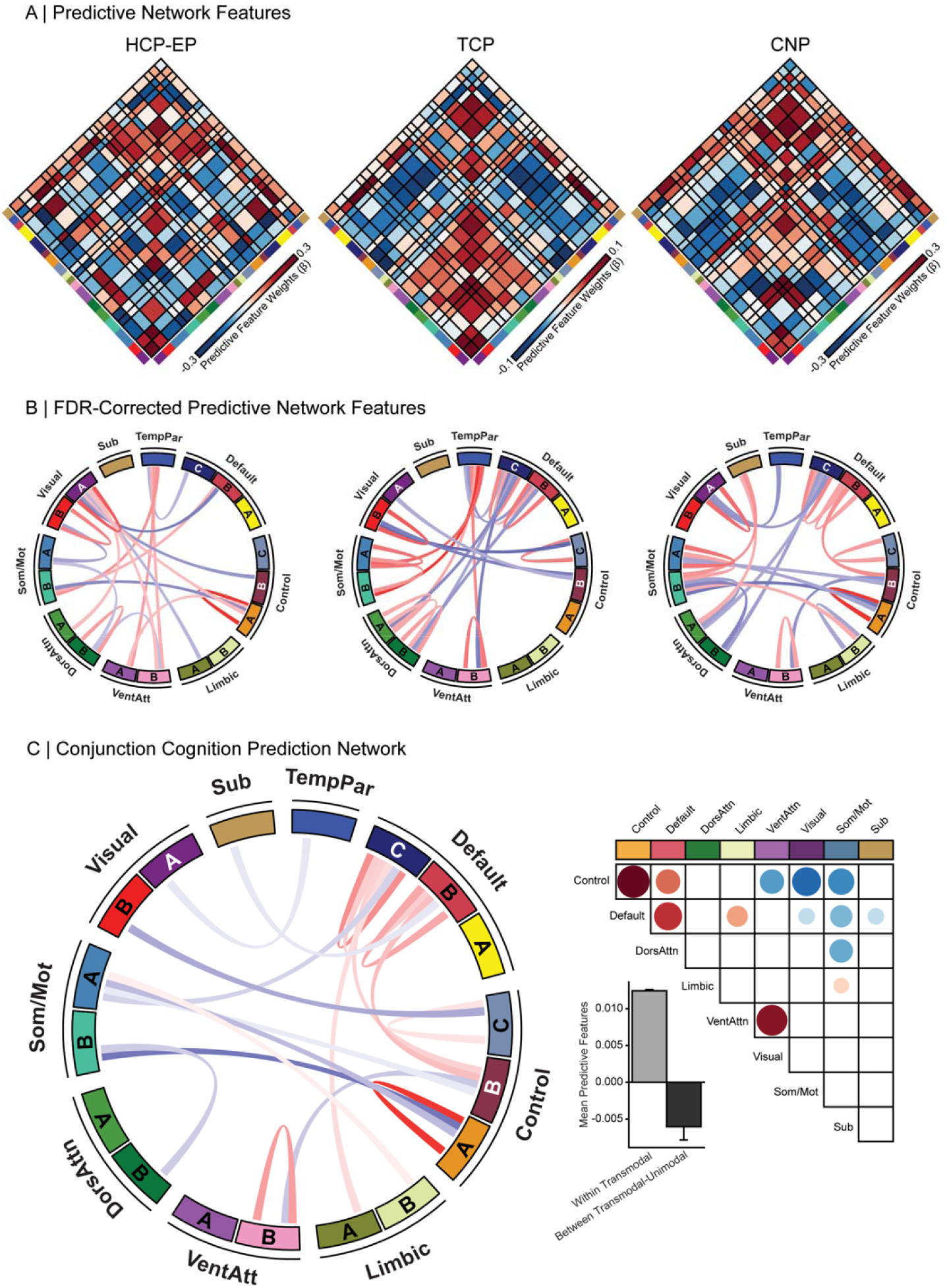
Increased within transmodal and reduced between network coupling is predictive of better cognitive functioning. **A)** Predictive feature matrices for each of the three datasets: Human Connectome Project – Early Psychosis (HCP-EP), Transdiagnostic Connectomes Project (TCP) & UCLA Consortium for Neuropsychiatric Phenomics (CNP), averaged within and between network blocks. Non-averaged data is provided in the supplement (FigS5). Red=positive predictive feature weight (stronger coupling predicts better cognition); blue = Negative predictive feature weight (weaker coupling predicts better cognition). **B)** Top 10% of FDR corrected predictive network connections for each dataset are displayed in Circos plots. **C)** Left: Circos plot showing the connections which survive multiple-comparison correction in a conjunction analysis across the three datasets. Top right: Heat map of conjunction analysis results aggregated into a 7-network and subcortex atlas solution. Bottom right: mean feature weights from the conjunction analysis categorized into within and between transmodal and unimodal networks. Sub = Subcortex; TempPar = Temporoparietal; DorsAttn = Dorsal Attention; VentAttn = Ventral Attention; SomMot = Somatomotor.

**Fig 4.**
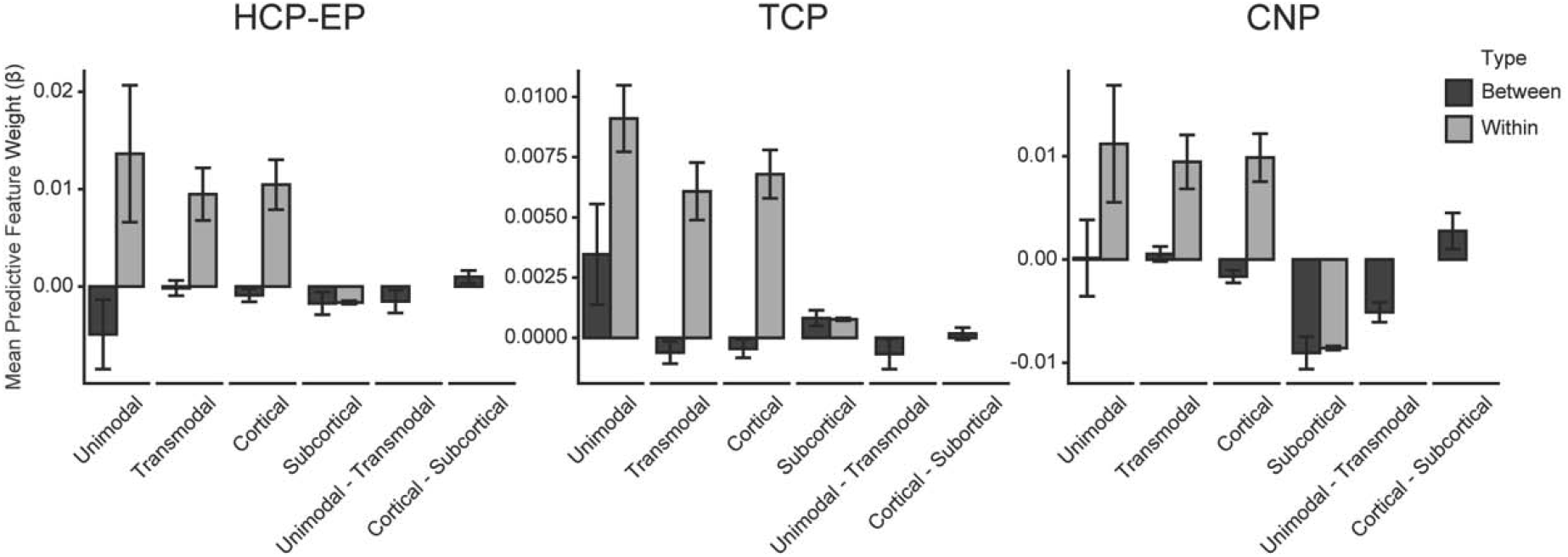
Increased within and decreased between system coupling predicts better cognition across datasets. Average predictive feature weights within (grey) and between (black) unimodal and transmodal cortical, subcortical, regions across the three datasets. Human Connectome Project – Early Psychosis (HCP-EP), Transdiagnostic Connectomes Project (TCP) & UCLA Consortium for Neuropsychiatric Phenomics (CNP). Error bars represent standard error around the mean. Unimodal networks include all visual and somatomotor networks, and transmodal networks include default, control, ventral attention, dorsal attention, limbic and temporoparietal networks.

To provide an increasing level of granularity, we also examined the network-level architecture of regional predictive features (Fig 5A). The strongest positive predictive regions for the HCP-EP dataset were the left cerebellar, right dorsal prefrontal and temporoparietal cortices and the negative predictive regions were right post-central and visual extrastriate cortices. For the TCP dataset, the strongest positive predictive regions included the right parahippocampal and left intra-parietal cortices and negative predictive regions included the right intra-parietal, anterior temporal and precuneus regions. For the CNP dataset, positive predictive regions included the bilateral hippocampus, right temporoparietal and dorsolateral prefrontal cortices and negative predictive regions were right post-central gyrus, somatomotor and left visual extrastriate cortex. While there was some heterogeneity in region-level predictive features, when these were aggregated into canonical networks (Fig 5B), across all datasets the strongest positive drivers of prediction performance were regions in transmodal temporoparietal, default, and frontoparietal networks. The strongest negative drivers also included the frontoparietal, dorsal attention, limbic and primary sensory regions, with the prominence of the frontoparietal network characterized by lower connectivity to sensory networks (Fig3C). We provide non-aggregated region-level distributions for each dataset individually, as well as distributions using a 7-network solution^51^ in the supplement (SFig5).

**Fig 5.**
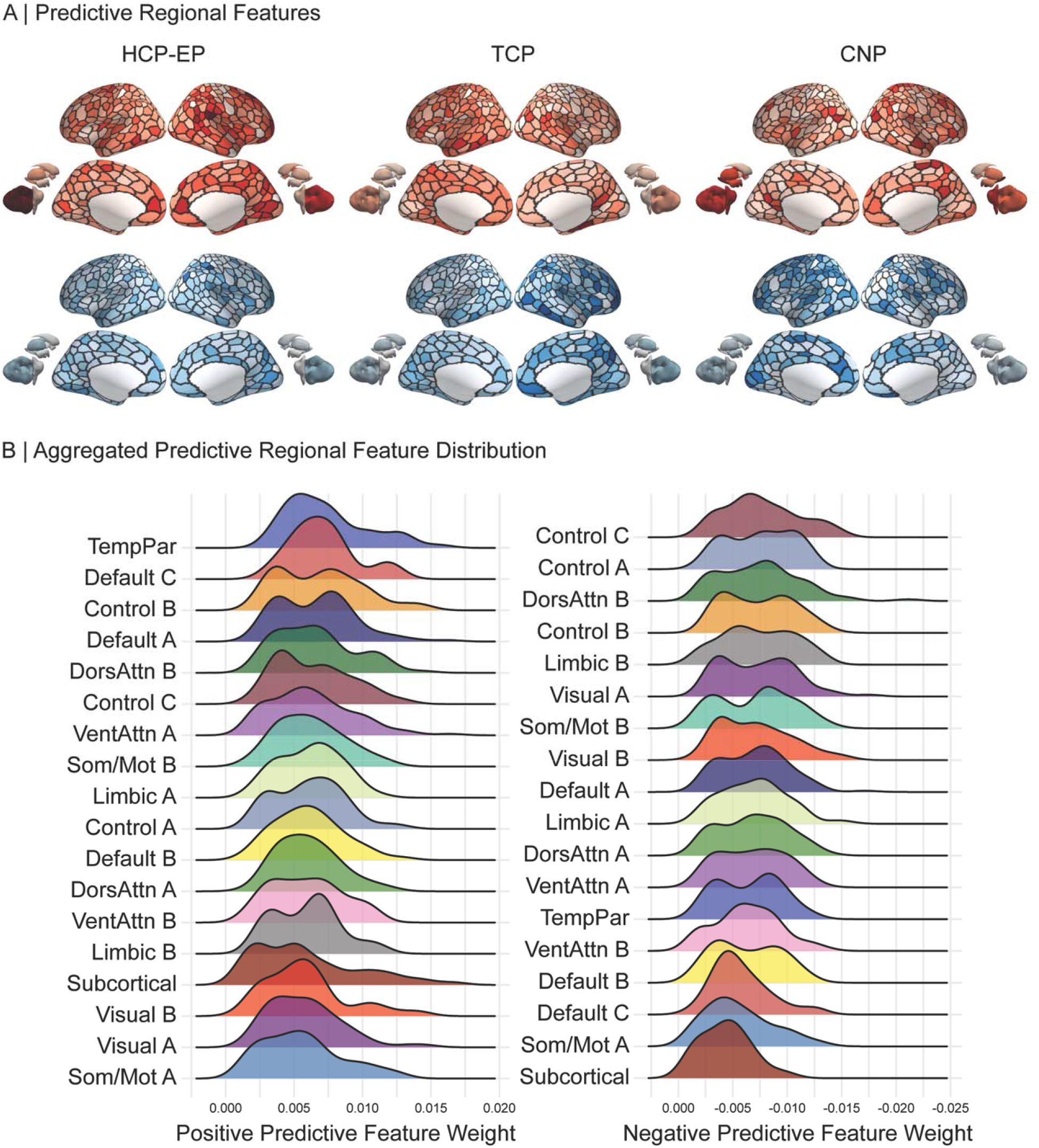
Predictive features at the regional-level. **A)** Regional feature weights projected onto cortical and sub-cortical regions. Average positive (red) and negative (blue) feature weights are shown separately for each of the three datasets. Human Connectome Project – Early Psychosis (HCP-EP), Transdiagnostic Connectomes Project (TCP) & UCLA Consortium for Neuropsychiatric Phenomics (CNP). **B)** Positive (left) and negative (right) distributions of regional feature weights from all three datasets aggregated into 17-networks and subcortex and ordered by strongest to weakest mean predictive feature weight.

## Discussion

Constructing robust models that reliably predict clinical symptoms from objective brain markers has previously required sample sizes exceeding most current collection efforts. Here we provide a proof of concept and define an associated roadmap for the generation of brain-based predictions in clinical populations. Critically, the models reported here are generalizable, maintaining prediction accuracy when trained in one dataset and tested on another, even when the datasets are independent and differ in their collection sites, demographic and diagnostic makeup, measures of global cognition, imaging acquisition sequences, and data-processing methods. The neurobiological features which drive prediction accuracy were most consistent between datasets at the scale of canonical functional networks rather than individual brain regions or edges. In line with previous hypotheses concerning the neurobiological substrates of cognition^14,15,19^, our findings converge on a global cognition predictive network where increased coupling within transmodal and the decreased coupling between transmodal and unimodal networks are linked with better cognition across transdiagnostic samples.

Widespread cognitive impairments are a core feature of common psychiatric illness, often presenting prior to illness onset^60,61^, and contribute to impaired social and occupational functioning^9–11^. Leveraging the meta-matching framework, we demonstrate that it is possible to achieve predictions of global cognitive functioning comparable to accuracies observed in the current state-of-the-art for the field, using sample sizes that are much smaller than those that have recently been recommended for deriving stable and generalizable brain-based predictions^38,40^. A particular advantage of our approach is that it yields discoveries that generalize across both healthy controls and common psychiatric disorders. By combining multivariate predictive models with transfer learning approaches like meta-matching, we provide a framework to leverage high-throughput population-based cohorts to boost predictive power in smaller datasets.

For a brain-based prediction model to have true clinical utility, it should be robust to sample characteristics, measurement methods, and data processing choices. This would allow a given approach to be deployed without overwhelming practical demands on the users, such as strict measurement and processing guidelines. Here we demonstrate that the meta-matching model generalizes not only across diagnostic categories, but also between independent datasets relying on different measures of cognition, neuroimaging protocols, and data processing strategies. Usually, models trained in one dataset lose much of their predictive capacity when applied to an independent dataset, even when the two datasets are diagnostically or demographically similar^2,38,41–43^. The meta-matching approach likely achieves this high level of generalizability by exploiting correlations amongst phenotypes, relying on a common set of neurobiological features which predict a broad range of behaviors that underlie an individual’s global cognitive performance, independent of diagnosis or measurement methods.

While we find commonality in neurobiological features driving prediction performance between the independent datasets at all spatial scales, the strongest correspondence was detected at the network level. Analogous to genetics, where broadening the spatial scale from single nucleotide polymorphisms to gene pathways results in more stable associations with complex behavior, we find that broadening the spatial scale from inter-regional edge-level connections to canonical networks results in stable associations. The similarity of neurobiological features at the network-level aligns with a large literature of explanatory studies implicating macro-scale networks as the primary unit-of analysis for complex behavioral traits^62,63^, as opposed to isolated regions or individual circuits, and evidence that the individual heterogeneity of patients assigned the same diagnosis is greatly attenuated when aggregating results across functional circuits and networks rather than brain regions^23^. Moreover, the consistency we observed between datasets suggests that the meta-matching model is likely making predictions by indexing a common neurobiology closely associated with cognitive function. In line with this hypothesis, we find that the connectivity of transmodal association networks, including the default mode and frontoparietal networks, is the most prominent driver of prediction accuracy. Specifically, increased connectivity within association networks, and decreased connectivity between these networks and visual and somatomotor sensory networks are associated with better cognition. This finding converges with decades of empirical work suggesting the activation and integrity of association networks is a critical driver of complex cognition^15,19,24^.

There are some limitations in the current work. While being able to make accurate and generalizable predictions of an observed phenotype such as cognition suggests the potential for clinical applications, future work should seek to develop models that can provide guidance on longitudinal outcomes. Such outcomes would include change in cognition, such as illness related decline, response to medications, and transitions to severe illness. As large-scale population based longitudinal data becomes available, the meta-matching framework can be adapted to predict symptom change and illness course, which could hold enormous clinical utility for psychiatry. Moreover, in our analyses we focused on global cognition, which can be reliably measured and is constantly impaired across common psychiatric diagnoses, and the assessment and treatment of cognitive changes closely align with patient goals. However, there are subcomponents of cognition which may be more impaired or less impaired across populations and future work should attempt to predict the results of more specialized neurocognitive assessments targeting constructs like working memory, processing speed or attention. Finally, in determining the features which are most relevant for predicting of cognition, we implemented the Haufe-transformation, which enhances both reliability and interpretability of feature weights. The transformation was initially designed for linear models and has not been extensively tested in non-linear and deep learning models. Nonetheless, we have previously demonstrated that the results of the transformation when using deep learning models in the prediction process are highly comparable to results using only linear models^47^.

By translating predictive models derived in large community-based datasets, we can make accurate and generalizable prediction of global cognition in transdiagnostic patient populations. The accuracy of these models is driven by increased coupling within transmodal networks and decreased coupling between transmodal and sensory networks.

## Methods

### Overview

Our overall analysis strategy aimed to develop a robust and generalizable model which can accurately predict global cognitive function in transdiagnostic patient samples. Briefly, we first used ‘meta-matching’^47^, which capitalizes on the correlation structure between phenotypes of clinical-interest and those available in larger population-level datasets by 1) training a Deep Neural Network (DNN) to predict a set of 67 health, behavioral and cognitive phenotypes using *in vivo* estimates of brain function from the UK biobank dataset^64^; 2) using this trained model to generate predictions of these phenotypes in smaller independent patient datasets; 3) and training and validating a prediction model which predicts global cognitive function using the predicted phenotypes generated from the DNN model in step 2. Global cognitive function was derived using principal components analysis on a range of neuropsychological tests that varied between the patient datasets. Significance of prediction performance and the generalizability of models was assessed using permutation testing, and the feature weights from each model were correlated between datasets and mapped at differing spatial scales (edge, region, network) to examine the consistency of neurobiological correlates.

### Datasets

This study used data from four datasets: the UK Biobank^64^, the Human Connectome Project Early Psychosis^65^, the Transdiagnostic Connectomes Project, and the Consortium for Neuropsychiatric Phenomics^66^. Our analyses were approved by the Yale University Institutional Review Board and the UK Biobank data was accessed under resource application 25163. Final number of included subjects, demographic and diagnostic characteristics are described below with additional details provided in Supplementary Table 1.

#### UK Biobank (UKBB)

The UK Biobank^64^ is a population epidemiology study of 500,000 adults aged 40–69 years and recruited between 2006 and 2010. A subset of 100,000 participants are being recruited for multimodal imaging, including brain structural MRI and rs-fMRI. A wide range of health, behavioral and cognitive phenotypes were collected for each participant. Here, we used the January 2020 release of 37,848 participants with complete and useable structural MRI and rs-fMRI.

#### Human Connectome Project Early-Psychosis (HCP-EP)

The HCP-EP^65^ study is acquiring high quality brain MRI, behavioral and cognitive measures in a cohort of people aged 16-35 years, with either affective or non-affective early phase psychosis within the first 5 years of the onset of psychotic symptoms. The dataset also includes healthy control participants and the data release used here (Release 1.1) comprises 140 patients and 63 controls. Inclusion and exclusion criteria for each respective dataset are described elsewhere^65^. In the current study, we used a subset of 145 participants who passed quality control and had complete and useable cognitive and rs-fMRI data. The included sample had a mean age of 23.41 (SD ± 3.68), was 38% female, and had a mean framewise displacement (head motion during rs-fMRI acquisition) of 0.06mm (SD ± 0.04).

#### Transdiagnostic Connectomes Project (TCP)

The TCP is an ongoing data-collection effort of to acquire brain MRI, behavioral and cognitive measures in a transdiagnostic cohort, including healthy controls and patients meeting diagnostic criteria for an affective or psychotic illness. Recruitment details, inclusion and exclusion criteria can be found in the Supplement. The data included in the current study was comprised of 101 participants who passed quality control and had complete and useable cognitive and rs-fMRI data, including 60 patients and 41 healthy controls. The included sample had a mean age of 32.21 (SD ± 12.54), was 57% female, and had a mean framewise displacement of 0.09mm (SD ± 0.05).

#### Consortium for Neuropsychiatric Phenomics (CNP)

The CNP dataset is a publicly available and comprised of brain MRI, behavioral and cognitive measures from 272 participants, including 130 healthy individuals and 142 patients diagnosed with affective, neurodevelopmental, or psychotic illnesses. Details about participant recruitment can be found elsewhere^66^. In the current study, we used a subset of 224 participants who passed quality control and had complete and useable cognitive and rs-fMRI data. The included sample had a mean age of 32.59 (SD ± 9.21), was 42% female, and had a mean framewise displacement of 0.08mm (SD ± 0.03).

### Quantifying Brain Function

#### MRI acquisition parameters

For the UK Biobank, a total of 490 functional volumes were acquired over 6 minutes at four imaging sites with harmonized Siemens 3T Skyra MRI scanners using the following parameters: repetition time = 735ms; echo time = 42ms; flip angle = 51°; resolution of 2.4mm^3^ and a multi-band acceleration factor of 8. For T1-weighted image, a MPRAGE sequence with a total of 256 slices were acquired using the following parameters: TR = 2000ms; TI = 880; resolution of 1mm^3^ and parallel imaging acceleration factor of 2.

For HCP-EP, a total of four runs of 420 functional volumes were acquired over 5.6 minutes at three imaging sites with harmonized Siemen 3T Prisma MRI scanners using the following parameters: repetition time = 800ms; echo time = 37ms; flip angle=52°; resolution of 2mm^3^ and a multi-band acceleration factor of 8. Spin-echo field maps in opposing acquisition direction were acquired to correct for susceptibility distortions. For T1-weighted image, a MPRAGE sequence with a total of 208 slices were acquired using the following parameters: TR = 2400ms; TI = 1000ms; and resolution of 0.8mm^3^.

For the TCP data, four runs of a total of 488 functional volumes were acquired over 5 minutes at two imaging sites with harmonized Siemen Magnetom 3T Prisma MRI scanners using the following parameters: repetition time = 800ms; echo time = 37ms; flip angle = 52°; resolution of 2mm^3^ and a multi-band acceleration factor of 8. Spin-echo field maps in opposing acquisition direction were acquired to correct for susceptibility distortions. For T1-weighted image, a MPRAGE sequence with a total of 208 slices were acquired using the following parameters: TR = 2400ms; and resolution of 0.8mm^3^.

For the CNP data, a total of 152 functional volumes were acquired over 5 minutes at 2 imaging sites with harmonized Siemen Trio 3T MRI scanners using the following parameters: repetition time = 2000ms; echo time = 30ms; flip angle = 90°; and a resolution of 4mm^3^. For T1-weighted image, a MPRAGE sequence with a total of 176 slices were acquired using the following parameters: TR = 1900ms; and resolution of 1mm^3^.

#### MRI Quality control

For all the clinical datasets, extensive quality control procedures were implemented, and the details can be found in the Supplement. Briefly, all raw images were first put through an automated quality control procedure (MRI-QC), which resulted in the exclusion of scans with large artifacts. Recent studies have shown that multiband datasets (i.e., HCP-EP, TCP) with high temporal resolution contain additional respiratory artifacts that manifest in the six realignment parameters typically used to calculate summary statistics of head motion^67,68^. To mitigate this effect, framewise displacement traces were downsampled and bandpass filtering was applied on realignment parameter between 0.2 and 0.5 Hz^69^. Following this step, uniform motion exclusion criteria were applied to all clinical datasets, using a previously established cut-off of mean framewise displacement greater than 0.55 which has previously been shown to result in good control of motion artifacts (Parkers et al., 2019). Finally, for all participants, functional connectivity matrices, carpet plots and QC-functional connectivity metrics were visualized and examined to ensure the processing and denoising steps achieved the desired effects of reducing noise and associations between head motion and functional connectivity.

#### MRI Processing

A detailed outline of the processing and denoising steps for each dataset is provided in the Supplement. Briefly, for each dataset we used differing but widely accepted processing strategies, which all included nonlinear spatial normalization to MNI space, brain tissue segmentation, and ICA-based denoising. These strategies were tailored to address differences in fMRI acquisition parameters (i.e., single- vs. multi-band) and to ensure that our models were robust to differences in preprocessing and denoising procedures. Global signal regression (GSR) was also applied to all scans, as we have previously demonstrated that it boosts prediction accuracy^70^ and improves data denoising^69,71^. The final derivatives used for prediction were 419□×□419 matrices for each subject, which were computed using 400 cortical and 19 non-cortical regions (for simplicity, non-cortical regions are indicated as ‘subcortex’; Fig1A) by averaging the time series within each parcel and computing a pair-wise Pearson correlation. For each subject, the correlation values were z-scored and the upper-triangle of this matrix which consisted of 87,571 unique functional connectivity estimates were entered into the prediction models.

### Quantifying Global Cognitive Functioning

For each of the three clinical datasets, principal component analysis was applied to all available cognitive and neuropsychological measures to derive a robust measure of global cognition. Each dataset had a distinct set of neuropsychological tests used to quantify cognitive functioning. A full list of measures for each dataset can be found in Supplementary Table 2. Briefly, for HCP-EP, measures included the National Institute of Health Toolbox^72^ and Wechsler Abbreviated Scale Intelligence^73^. For TCP, measures were administered online through the TestMyBrain platform^74^ which included assessment of matrix reasoning, sustained attention, basic psychomotor speed and processing speed, as well as Stroop and Hammer reaction time measures acquired during MRI acquisition. For the CNP, measures included subtests from California Verbal Learning Test, Wechsler Memory Scale^75^ and Wechsler Adult Intelligence Scale^76^. To reduce model complexity, the PCA for each dataset was computed on the full sample, before any cross-validation. To ensure that data-leakage between the train and test splits did not influence our results, we tested whether prediction models generalized between the different and completely independent datasets (see *Evaluating Model Generalizability*). For each dataset, the first principal component (PC) was retained. For the HCP-EP dataset the first PC explained 57.2% of the variance, with the second and third PC examining 13.6% and 6% of the variance respectively. For the TCP dataset the first PC explained 25.9% of the variance, with the second and third PC examining 16.2% and 13.8% of the variance respectively. For the CNP dataset the first PC explained 32.5% of the variance, with the second and third PC examining 9.4% and 7% of the variance respectively. For each dataset, a higher PC score indexed better global cognition. The full list of loadings for each item can be found Supplementary Table 2.

### Brain-based Predictive Modelling

Consistent with the approach outlined by He, et al. ^47^, we trained a single fully-connected feedforward Deep Neural Network (DNN) using the UK Biobank dataset to predict 67 different cognitive, health and behavioral phenotypes from resting-state functional connectivity matrices. This type of DNN possesses a generic architecture, where the connectivity values enter the model through an input layer, and each other layer is fully connected to the layer before it, meaning that values at each node are the weighted sum of node values from the previous layer. During the training process, these weights are optimized so that the output layer results in predictions that are close estimations of observed phenotypes. The 67 cognitive, health and behavioral variables were selected based on an initial list of 3,937 phenotypes by a systematic procedure that excluded brain variables, categorical variables (except sex), repeated measures and phenotypes that were not predictable using a held-out set of 1,000 participants^47^. A full list of selected phenotypes can be found in supplementary Table 3 and further details of the DNN architecture and variable selection procedure within the UK Biobank can be found elsewhere^47^. This trained DNN model is openly available and can be implemented in any sample with available resting-state functional connectivity data (see https://github.com/ThomasYeoLab/Meta_matching_models).

Following training of the DNN, it was applied to the clinical datasets using a nested cross-validation and stacking procedure. The procedure described below was implemented separately for each clinical dataset. First, the DNN was applied to the dataset, using resting-state functional connectivity matrices as inputs, resulting in 67 generated cognitive, health and behavioral variables as outputs. These outputs and corresponding global cognitive scores were split into 100 distinct train (70%) and test (30%) sets without replacement. We then implemented a stacking procedure, where a Kernel Ridge Regression (KRR) model using a linear kernel with l2-regularization was trained to predict global cognitive functioning scores using the generated 67 cognitive, health and behavioral variables as inputs. KRR is a classical machine learning technique that makes a prediction of a given phenotype in an individual as a weighted version of similar individuals. Similarity was defined as the inter-individual correlation of predicted phenotypes. KRR has one free parameter which controls the strength of regularization and was selected based on 5-fold cross-validation within the training set. Once optimized, the model was evaluated on the held-out test set. This was repeated for the 100 distinct train-test splits to obtain a distribution of performance metrics.

As a comparison to the meta-matching model described above, we also implemented a standard machine learning model to provide a baseline. Here we used the standard implementation of KRR, where the model was trained to predict global cognitive function scores, using resting-state functional connectivity matrices as inputs. This is in contrast to the KRR model implemented during the meta-matching stacking process, which was trained using the DNN-generated cognitive, health and behavioral as inputs. The nested cross-validation procedure used for the baseline comparison model was the same as the one used for the meta-matching model, where each dataset was split into 100 distinct train (70%) and test (30%) sets without replacement, followed by 5-fold cross-validation within the training set to tune the model hyperparameters and the model performance was evaluation on the held-out test set. All code used for analysis and figure generation can be found on-line at https://github.com/sidchop/PredictingCognition.

### Evaluating Model Accuracy

The accuracy of each model is defined as the Pearson correlation between the true and predicted behavioral scores for each split. Average accuracy was computed by taking the mean across the 100 distinct train-test splits. We also evaluated absolute, as opposed to relative, prediction accuracy using the coefficient of determination (SFig2; ^1^). All models were evaluated on whether they performed better than chance using null distributions of performance metrics. For the meta-matching model, in each of the three clinical datasets, cognitive function scores were randomly permuted 10,000 times. Each permutation was used to train (70% of sample) and test (30% of sample) a null prediction model. The p-value for the model’s significance was defined as the proportion of null prediction accuracies greater than the mean performance of the observed model. The same procedure was used to evaluate statistical significance of the baseline comparison model.

### Evaluating Model Generalizability

The generalizability of the model was evaluated by training the meta-matching model on all individuals from one dataset and testing on all individuals from another dataset. This results in six train-test pairs between the three clinical datasets (i.e., HCP-EP, TCP and CNP). For each train-test pair, accuracy was again measured as the Pearson correlation between the predicted and actual scores on the test dataset. We also report absolute accuracy using the coefficient of determination (SFig2; ^1^). Statistical significance was evaluated by permuting the training dataset cognitive function scores and computing a null meta-matching model 10,000 times. The p-value corresponding to model significance was defined as the proportion null prediction accuracies greater than the performance of the observed model. To compare the within dataset prediction performance between standard baseline comparison and meta-matching models, we computed a paired-sampled t-test for each of the three clinical datasets.

### Comparing Neurobiological Features Between Datasets and Spatial Scales

To increase the interpretability and reliability of feature weights from the prediction models, we used the Haufe transformation^52–54^ which considers the covariation between the training set functional connectivity and global cognition scores. This procedure ensures that the feature weights index quantities that are statistically related to global cognition and results in a positive or negative predictive feature value for each edge of the functional connectivity matrix. A positive predictive feature value indicates that higher functional connectivity for the edge was associated with the greater predicted cognitive functioning and a negative predictive feature value indicates that lower functional connectivity was associated with the greater predicted cognitive functioning. For each of the three models, the transformed feature weights were then averaged across the 100 splits to obtain mean feature weights, resulting in a single symmetric 419 × 419 predictive feature matrix for each dataset.

We assessed the association of neurobiological predictive features between each of the three predictive feature matrices at the edge-, region-, network-level. At the edge-level, which comprises each of the 87,571 feature weights, similarity was assessed using Pearson correlation. To account for spatial autocorrelation between each pair of feature weight matrices^55^, we applied the spin-test, where the cortical regions of the atlas are rotated on the inflated surfaces to generate 10,000 null atlas configurations which preserve the spatial autocorrelation pattern of the cortex. These null atlas configurations are used to shuffle the rows and columns of the feature weight matrices, allowing the generation of a null distribution of Pearson correlation values between a pair of feature weight matrices at the edge-, region- and network-level. Statistical significance was assessed as the proportion of null values greater than the observed value (p_*spin*_). As the spin-test procedure can only be applied to cortical regions, the 19 non-cortical regions were excluded when computing the p-value. By taking the mean of all edges attached to each of the 419 brain regions, edges-level connections can be aggregated into region-level predictive features. By taking the mean of all edges within and between 18 canonical functional networks including the subcortex Fig1A; ^48^, edges-level connections can also be aggregated into 171 network-level predictive features. For both aggregated scales (region- and network-level), positive and negative feature weights were considered separately by zeroing negative or positive values before averaging, respectively. To compare the negative and positive feature weight correlations within each spatial scale, we used Fishers Z-statistic modified for non-overlapping correlations based on dependent groups^77,78^.

### Evaluating Neurobiological Features

To evaluate the statistical significance of feature weights for each of the three datasets, we implemented a permutation testing procedure. To reduce the multiple comparison burden, we evaluated the significance of each model at the network-level, where the observed feature weights for each model were averaged within and between 18 network blocks, resulting in 171 network-level features per model. This network averaging procedure was repeated for feature weights from 2000 null models, where the cognitive function score had been randomly permuted. This results in null-distribution of network-level feature weights for each of the 171 network connections and the p-value was computed as the proportion of the null network-level feature weights greater than the observed value. The p-values were then FDR corrected and evaluated at a *p* < 0.05 level. To uncover the network-level predictive features which drive accuracy across the three datasets, we implemented a conjunction analysis, where at each network connection, the minimum FDR corrected p-value was retained, and evaluated for significance at a *p* < 0.016, accounting for the three contrasts.

### Control Analyses

Demographic characteristics such as age and sex, as well as head motion during neuroimaging can bias the performance of prediction models^79^. To ensure that the model performance was not driven by these covariates, we repeated the primary models after adjusting for age, sex and mean framewise displacement. For each of the 100 train-test splits, the variables were first regressed out of the global cognition training data, and the resulting regression coefficients were used to residualize the global cognition test data^80^. The reported results were robust to covariate inclusion and all three meta-matching models remained statistically significant (FigS6; FigS7). Moreover, the edge-level feature weights from the original and covariate adjusted models were highly correlated at *rs* > 0.96 for all three datasets (FigS7). Performance remained stable in the HCPEP sample (*r* = 0.51) and decreased in the TCP (*r* = 0.25) and CNP datasets (*r* = 0.28; Fig S6). The pattern of results showing superior performance of the meta-matching compared to conventional KRR model was maintained in all three datasets (FigS6).

Meta-matching capitalizes on correlations between neurobiology associated with diverse demographic, health, and behavioral phenotypes. By examining the feature weights associated with the DNN-generated demographic, health, and behavioral phenotypes that are computed during the stacking part of the meta-matching model, it is possible to evaluate which generated demographic, health, and behavioral phenotypes are driving prediction of cognitive outcomes. The generated variables driving performance were highly consistent across the three datasets (*all rs* > 0.95; FigS8). The primary drivers of prediction were directly related to cognition (fluid intelligence, matrix pattern completion, symbol digit substitution). However, across the three datasets, both age and the first genetic principal component were strong predictors, with the latter indexing ancestry, which can also be a proxy for complex forms of societal and environmental bias. To investigate whether the observed improvements in behavior prediction performance extend beyond the specific demographic factors that can encapsulate societal bias (ancestry, sex, and age), in turn affecting cognitive performance, we repeated the meta-matching stacking procedure for our primary analyses after removing the first genetic principal component, age and sex from the meta-matching model and found near identical results to our original model (FigS9).

## Data Availability

All data produced in the present study are available upon reasonable request to the authors.

https://github.com/sidchop/PredictingCognition

## Acknowledgements

This work was supported by the National Institute of Mental Health (R01MH123245 to AJH and R01MH120080 to AJH and BTTY) and the Kavli Institute for Neuroscience at Yale University (Postdoctoral Fellowship for Academic Diversity to ED. AF was supported by the Sylvia and Charles Viertel Foundaton, National Health and Medical Research Council (IDs: 1197431 and 1146292) and Australian Research Council (IDs: DP200103509). BTTY is supported by the Singapore National Research Foundation (NRF) Fellowship (Class of 2017), the NUS Yong Loo Lin School of Medicine (NUHSRO/2020/124/TMR/LOA), the Singapore National Medical Research Council (NMRC) LCG (OFLCG19May-0035), and NMRC STaR (STaR20nov-0003). KA is a scientific advisor to and shareholder in BrainKey Inc., a medical image analysis software company. ERO is supported by an American Australian Association Graduate Education Fund Scholarship.

## Supplement

**Supplementary Table 1.** Sample demographics

|  | **HCP-EP**<br>(n=145) | **TCP**<br>(n=101) | **CNP**<br>(n=224) |
| --- | --- | --- | --- |
| **Age** (mean,sd) | 23.41(3.86) | 32.21(12.54) | 32.59(9.21) |
| **Sex** (f, %) | 57, 38% | 50, 57% | 95, 42% |
| **Head motion** (mm, sd) | .06(.04) | .09(.05) | .08(.03) |
| **Diagnosis** |  |  |  |
| *SZ\** | 62 | 4 | 37 |
| *SZAD* | 8 | 2 | 0 |
| *MDD* | 5 | 19 | 0 |
| *BD* | 20 | 6 | 40 |
| *ANX\** | 0 | 5 | 0 |
| *ADHD* | 0 | 0 | 37 |
| *OCD* | 0 | 0 | 0 |
| *PTSD* | 0 | 6 | 0 |
| *SUD* | 1 | 2 | 0 |
| *ED* | 0 | 2 | 0 |
| *None (HC)* | 52 | 41 | 110 |

Acronyms: HCP-EP = Human Connectome Project – Early Psychosis, TCP = Transdiagnostic Connectomes Project, CNP = UCLA Consortium for Neuropsychiatric Phenomics, SZ = Schizophrenia, SZAD = Schizoaffective Disorder, MDD = Major Depressive Disorder, BD = Bipolar Disorder, ANX=Anxiety Disorder, ADHD = Attention Deficit Hyperactivity Disorder, OCD = obsessive Compulsive Disorder, PTSD = Post-Traumatic Stress Disorder, SUD = Substance Use Disorder, HC = Healthy Control.
*\*Includes all Schizophrenia Spectrum Diagnosis, except Schizoaffective Disorder*
*\*Includes Generalized Anxiety Disorder and Specific Phobia*

**Supplementary Table 2.**
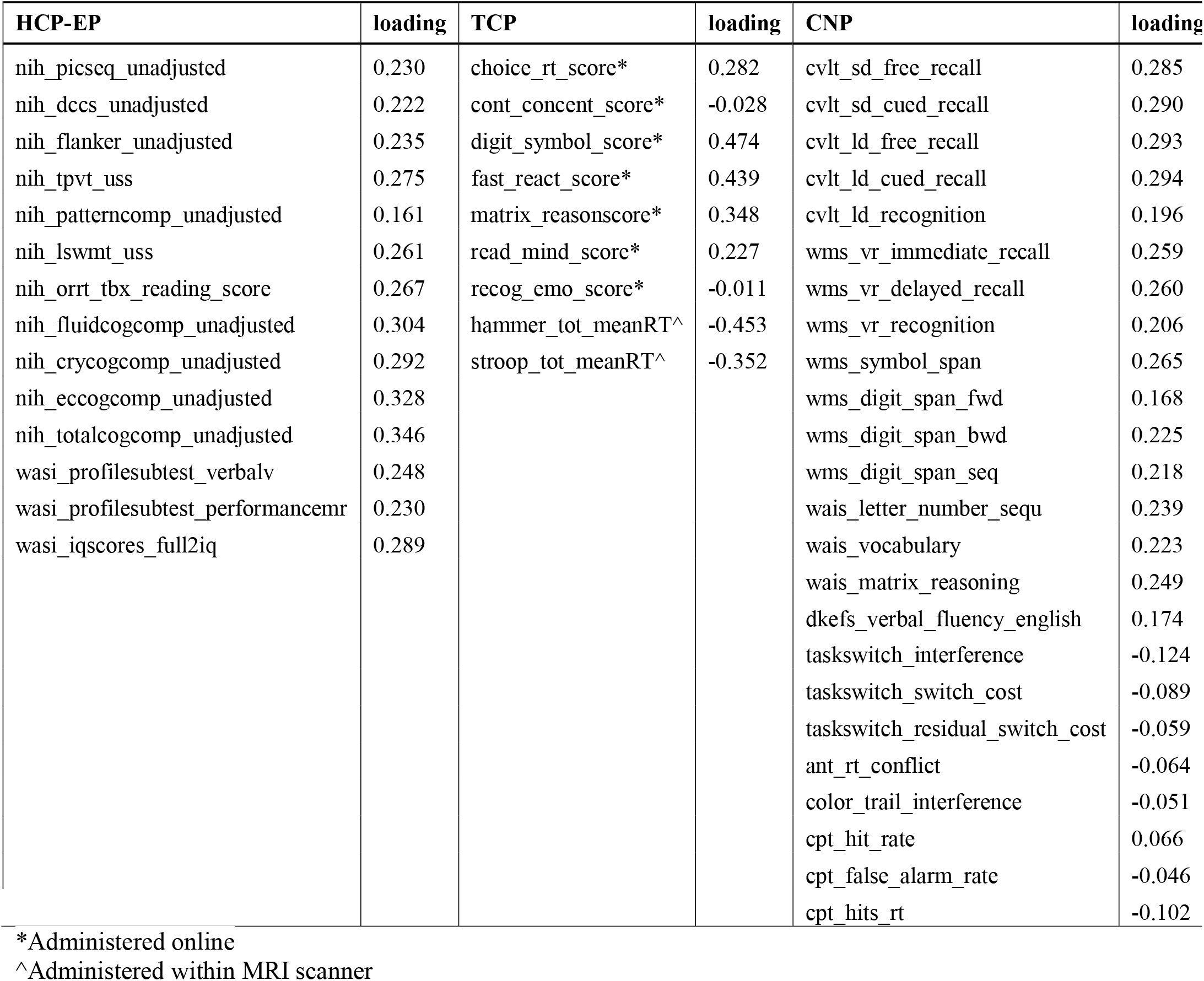
Cognitive tests for each clinical dataset and loadings on the first principal component.

**Supplement Table 3.**
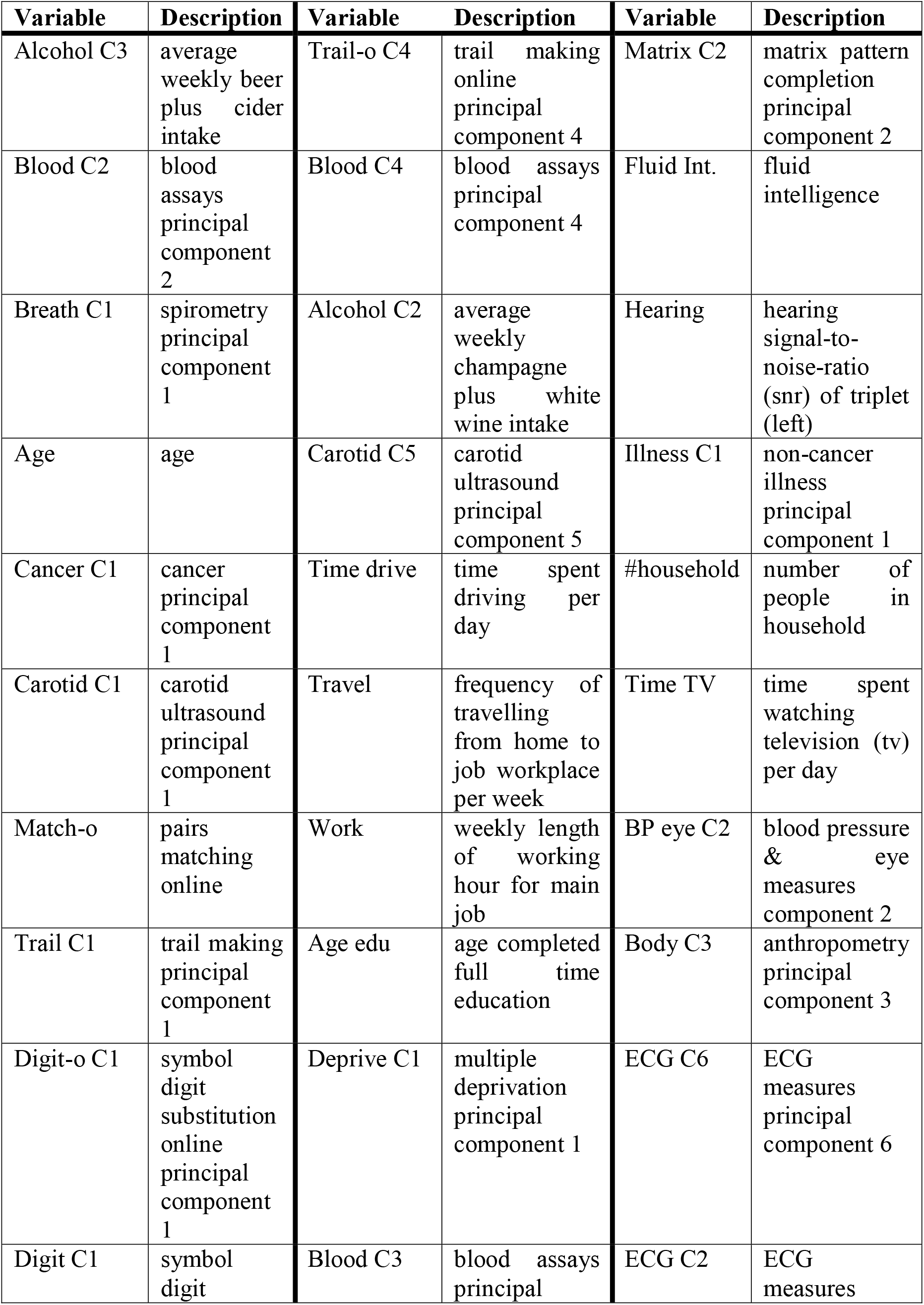
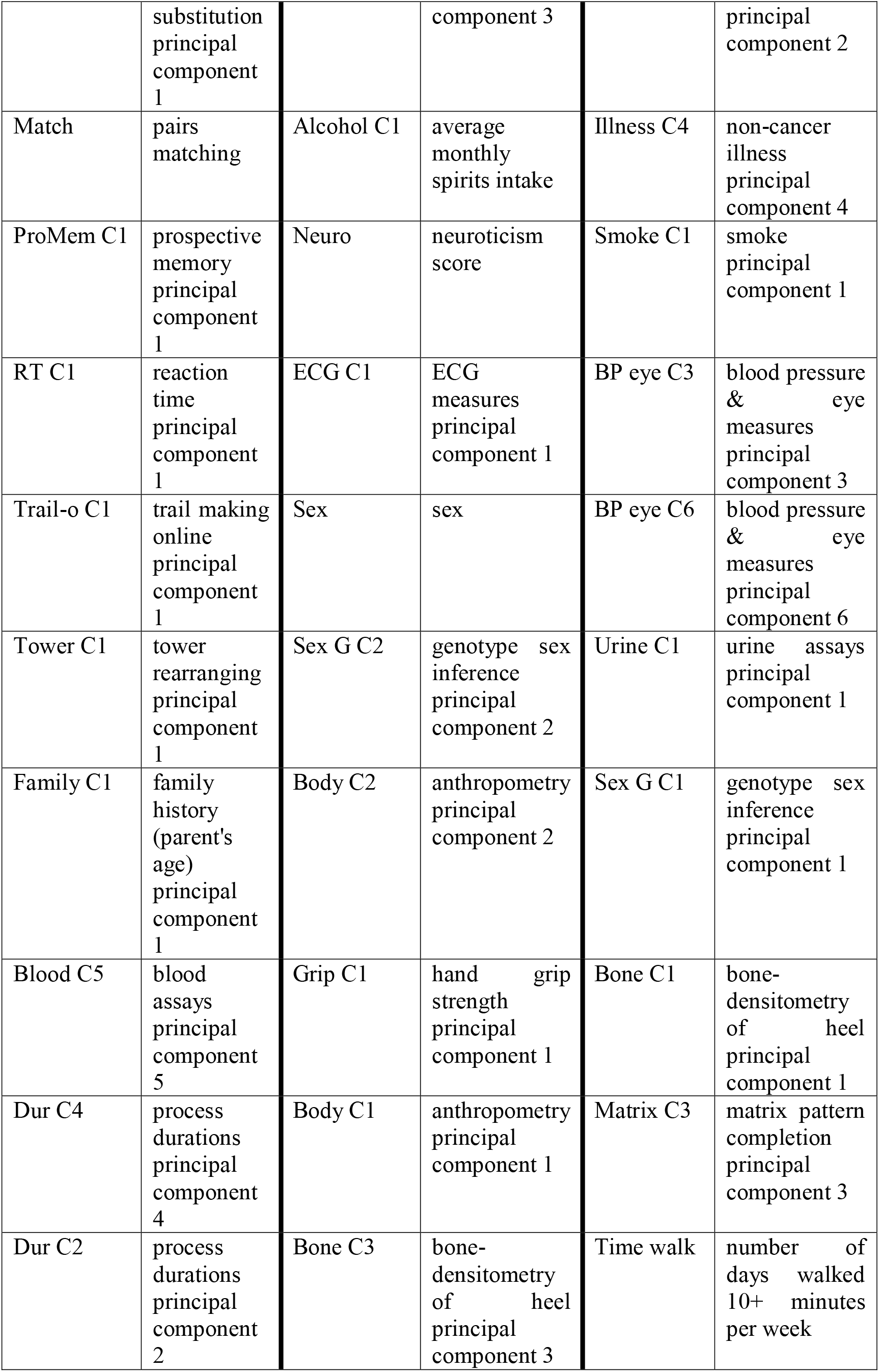
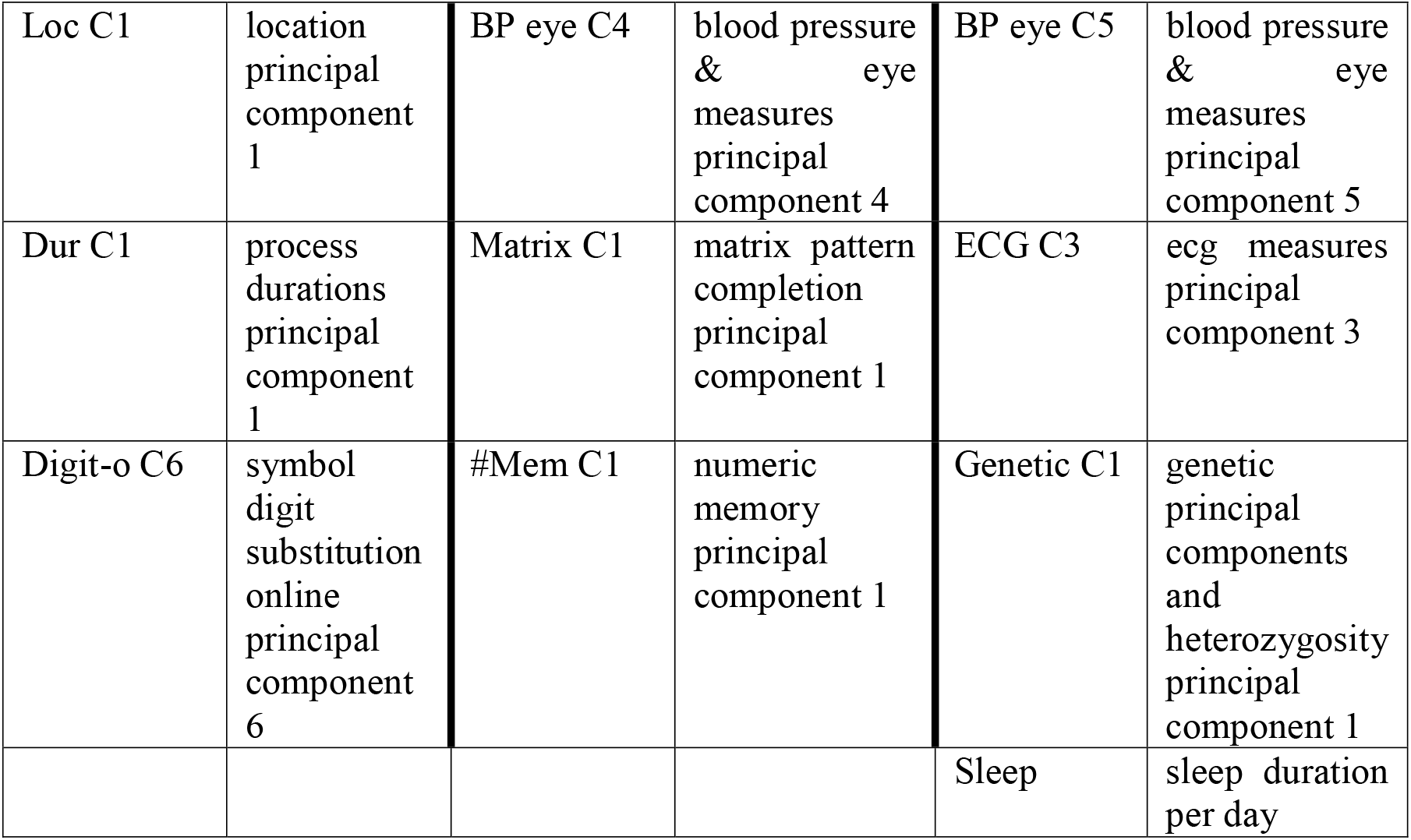
67 Phenotypes from Biobank used to train meta-matching model

**Supplementary Figure 1.**
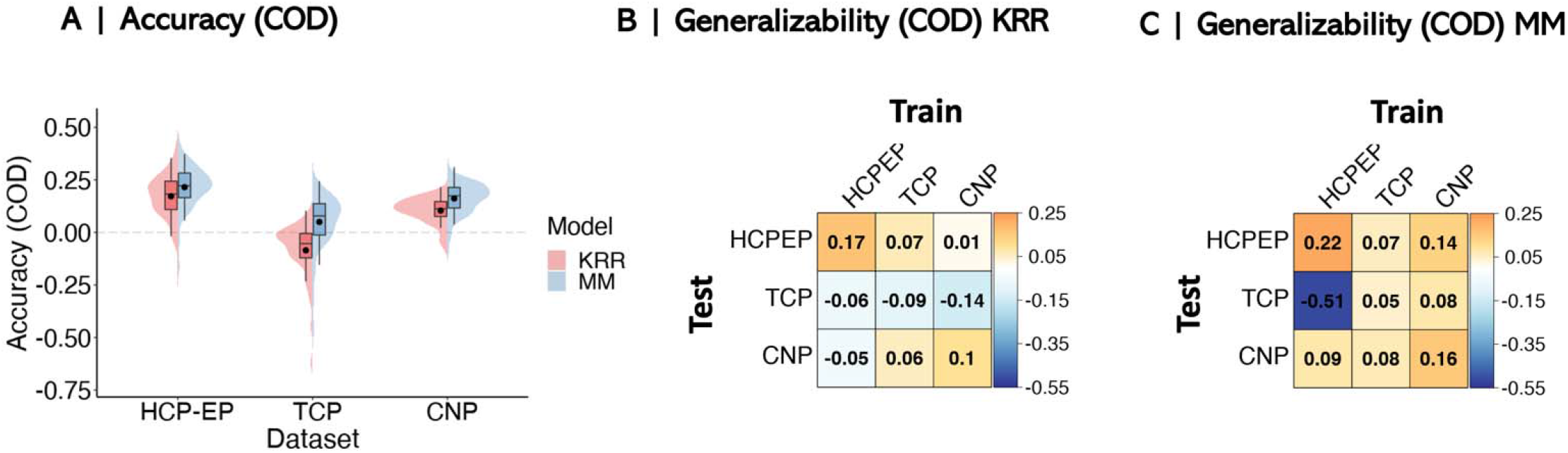
Model accuracy and generalizability assessed using Coefficient of Determination (COD)

**Supplementary Figure 2.**
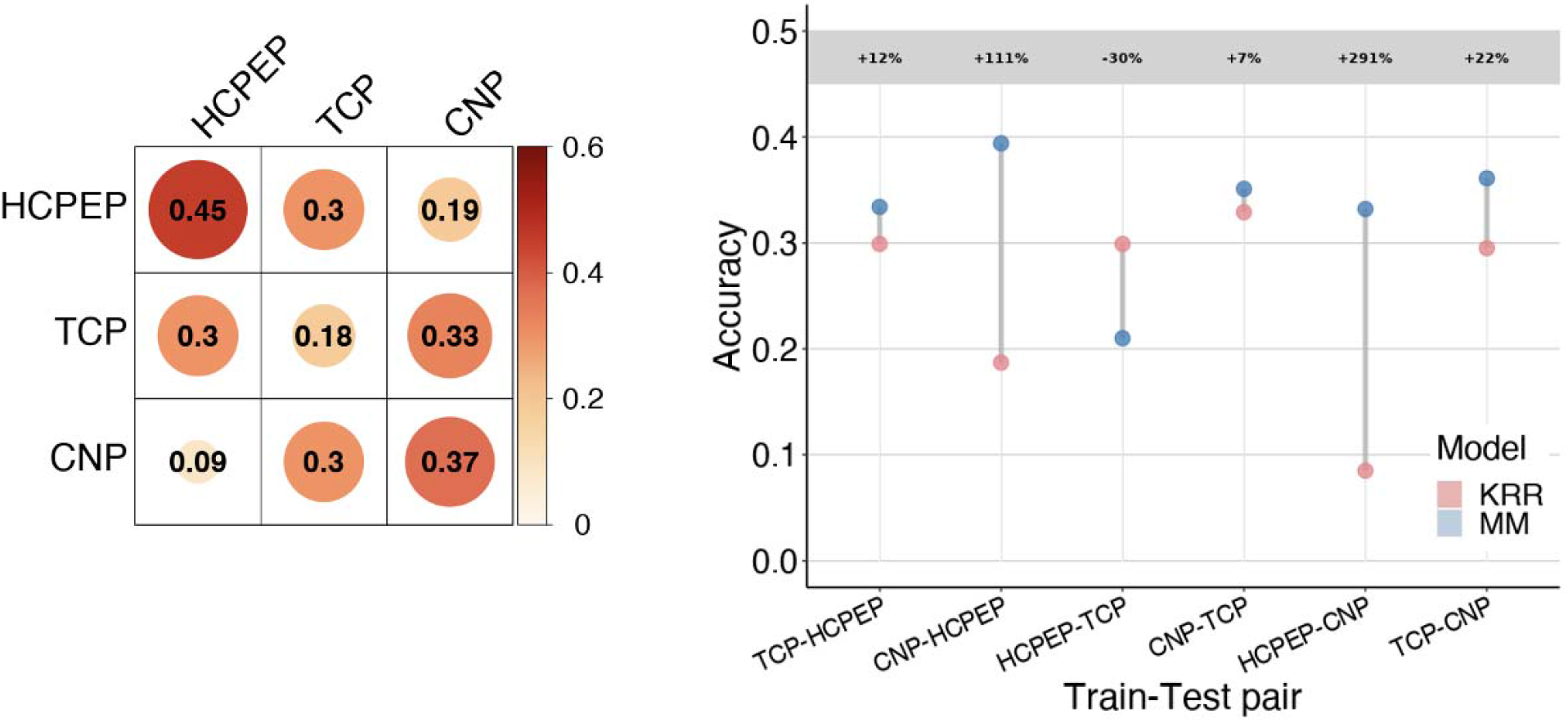
Kernel Ridge Regression (KRR) model generalizability matrix (left) and differences in generalizability between KRR and meta-matching (MM) models (right).

**Supplementary Figure 3.**
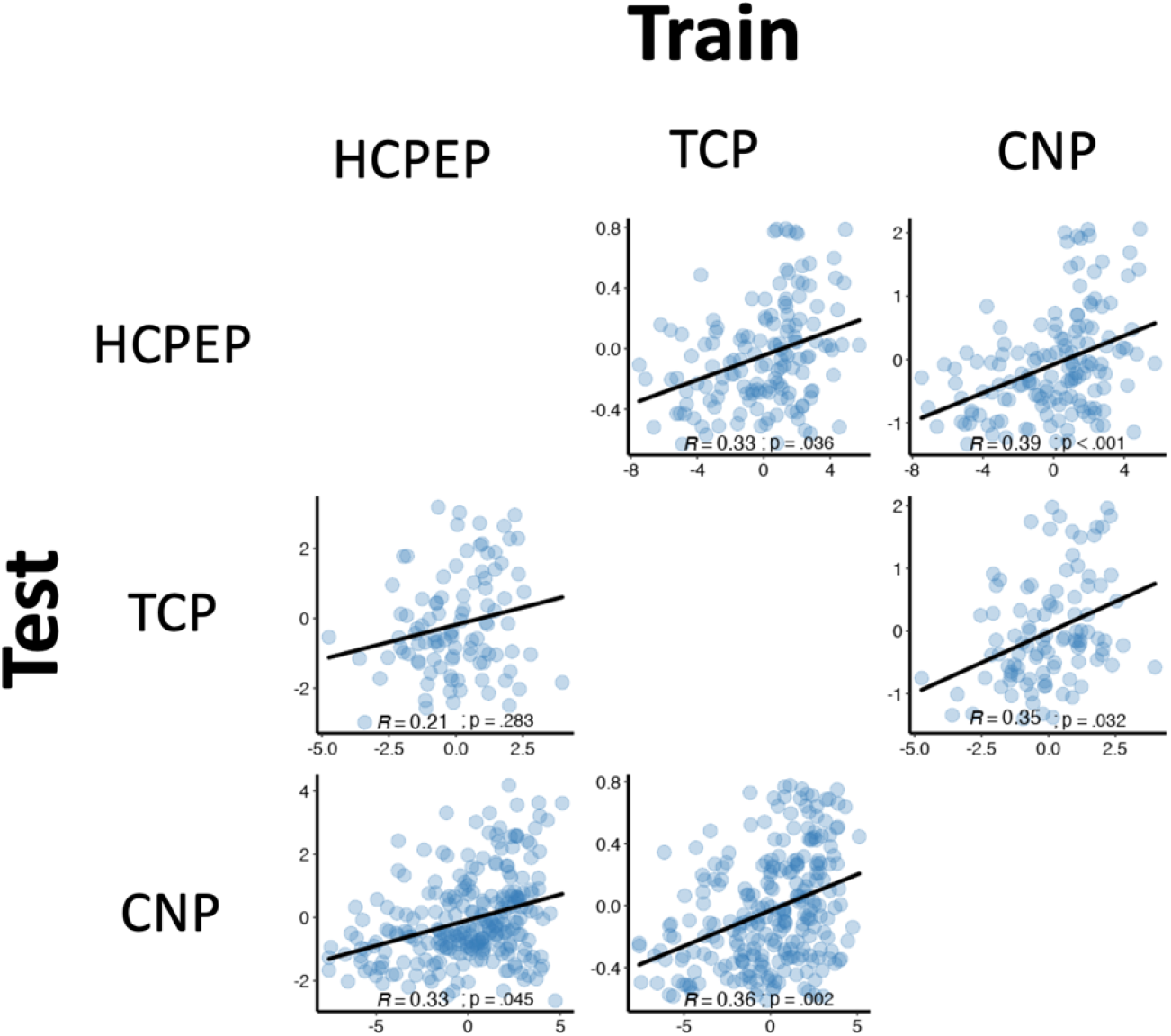
Scatterplots of observed and predicted cognition scores for generalizability of the meta-matching model.

**Supplementary Figure 4.**
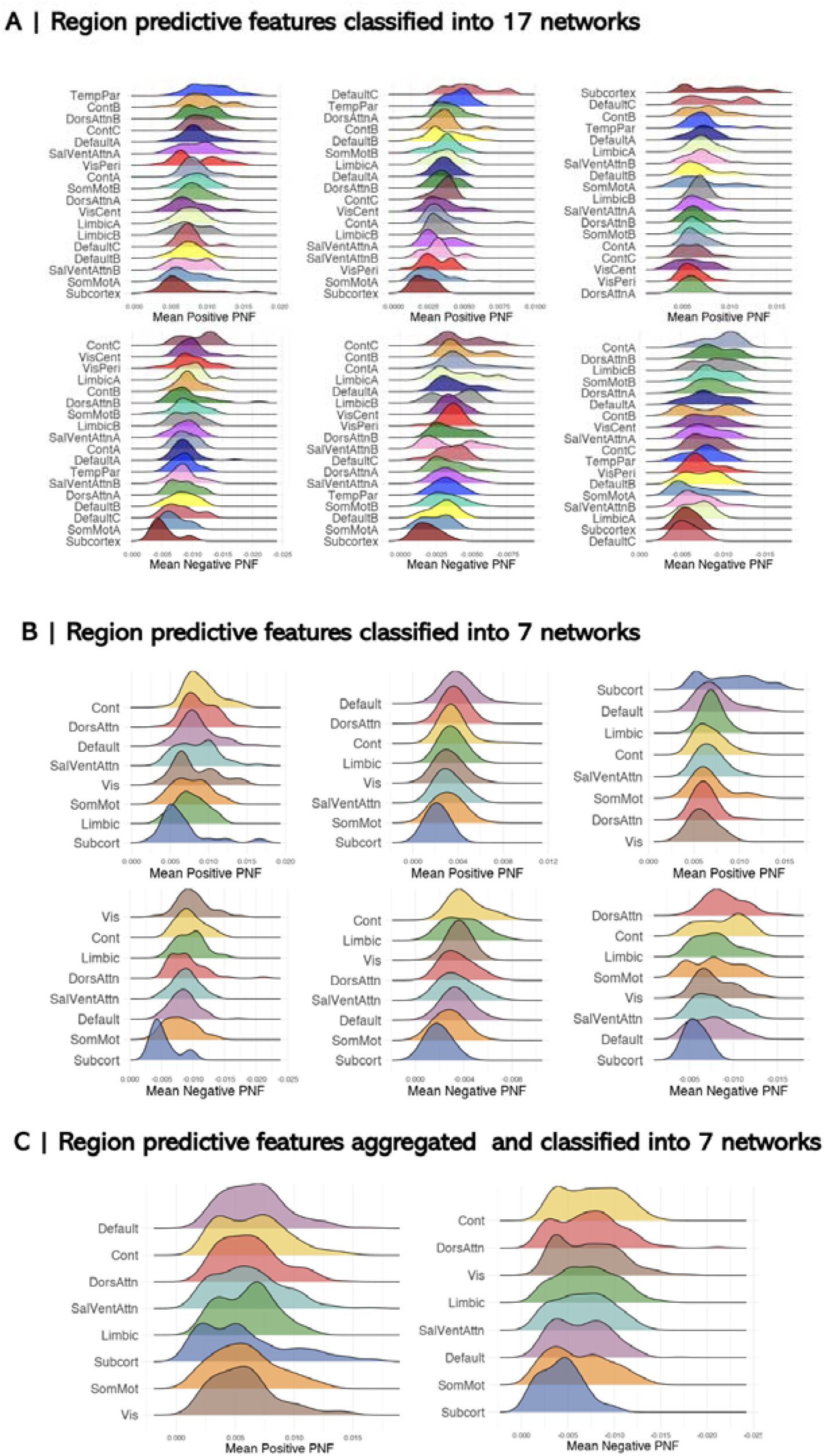
Regional predictive features classified into 7 and 17 network solutions.

**Supplementary Figure 5.**
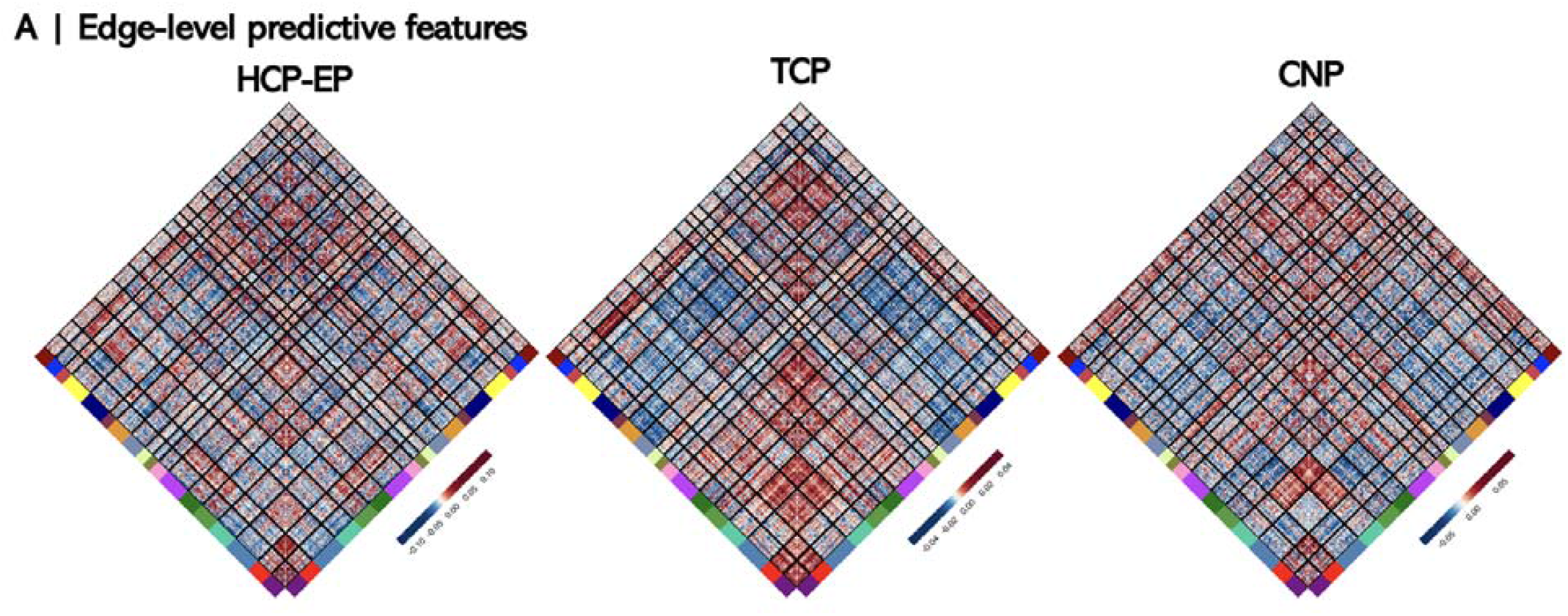
Edge-level predictive feature weights for each dataset

**Supplementary Figure 6.**
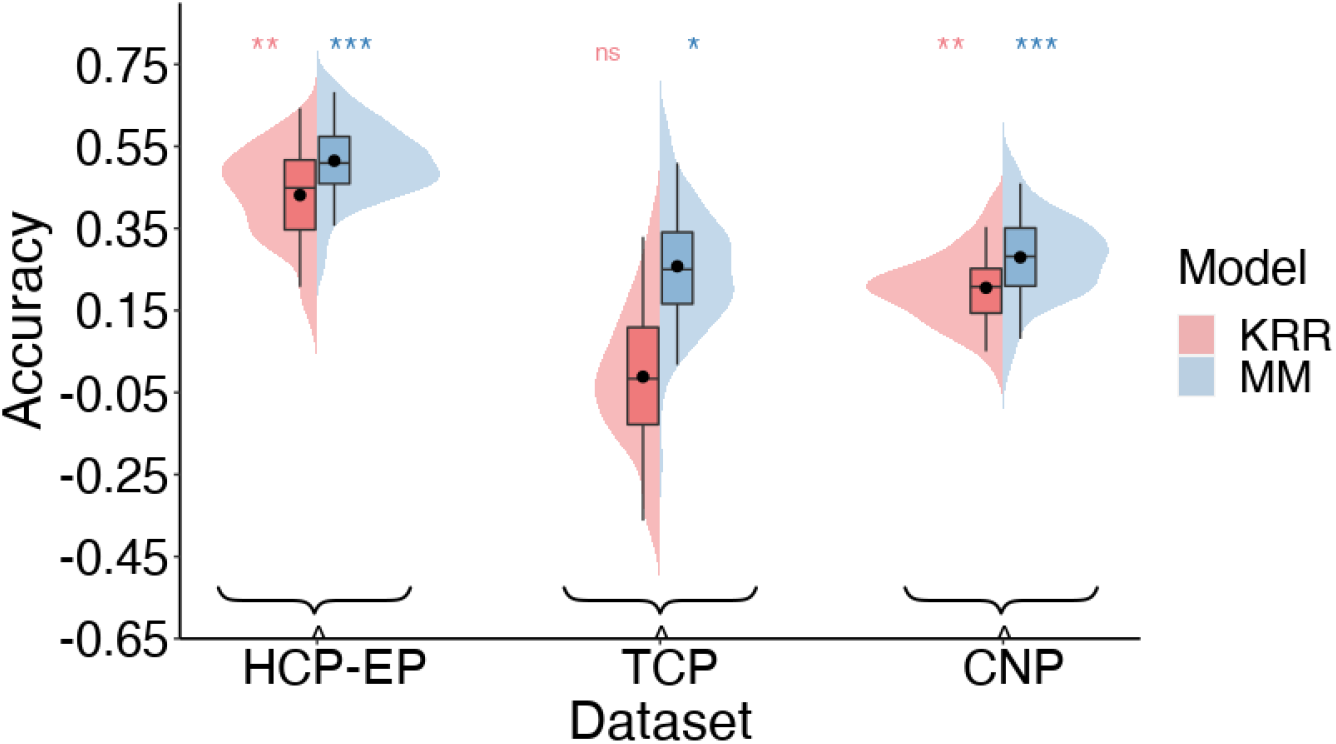
Model performance after regressing out age, sex and head motion (mean FD)

**Supplementary Figure 7.**
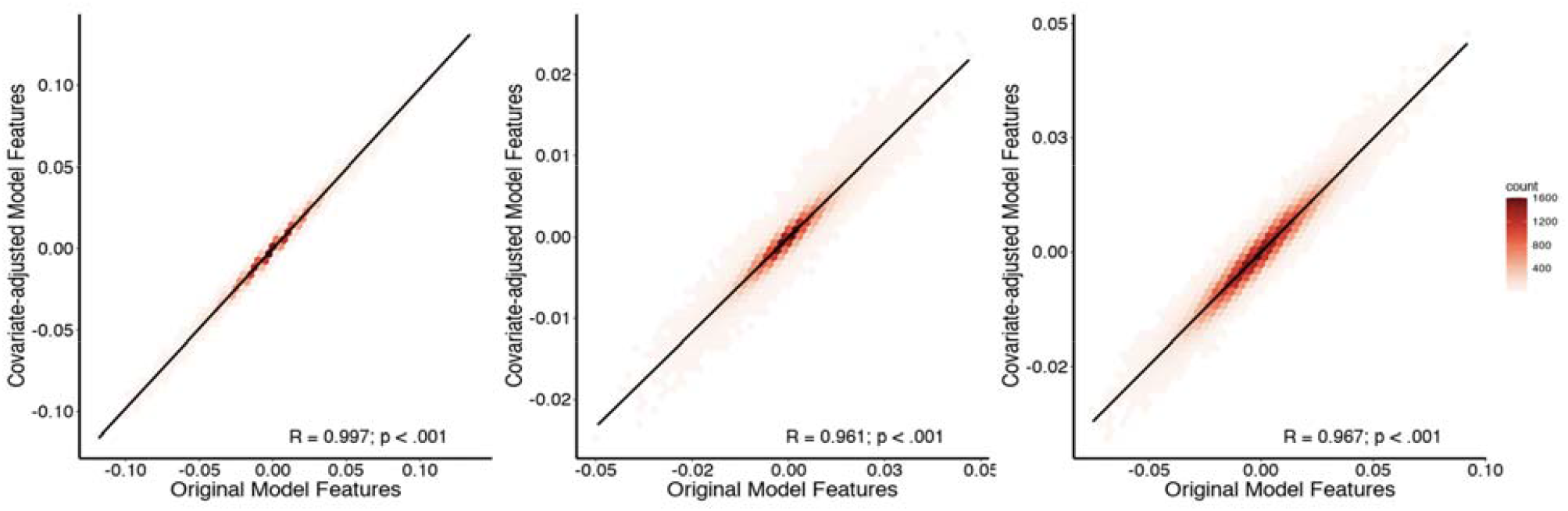
Correlation between edge-level feature weights for original and covariate adjusted meta-matching models.

**Supplementary Figure 8.**
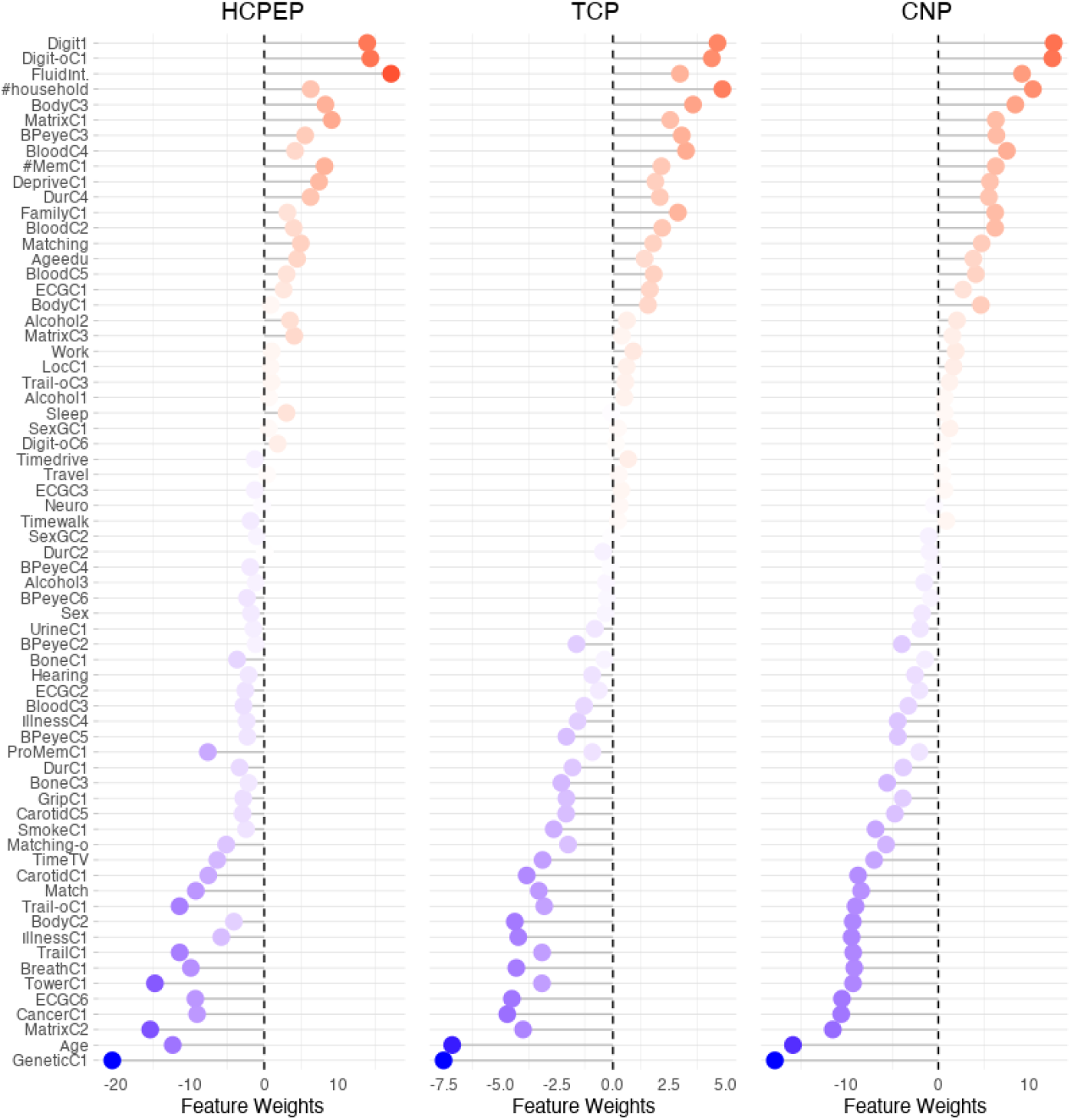
Feature weights associated with 67 health, demographic and behavioral variables using the stacking component of the meta-matching model.

**Supplementary Figure 9.**
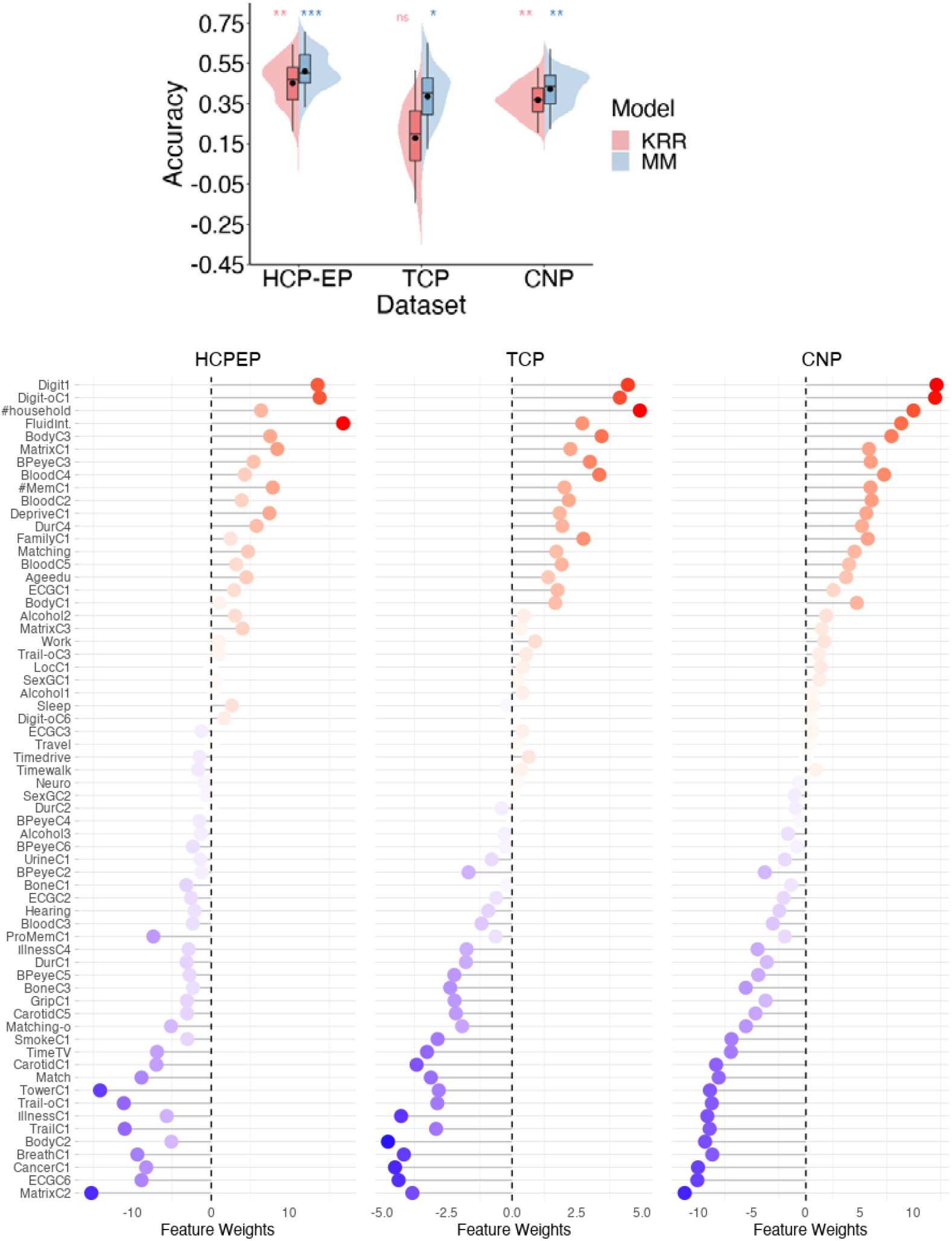
Model performance (Top) and feature weights (bottom) associated with 64 health, demographic and behavioral variables using the stacking component of the meta-matching model (after removing age, sex, and gene PC1 from the meta-matching model).

### Additional Information on TCP dataset

The Transdiagnostic Connectome Project is ongoing data-collection effort between Yale University and McLean Hospital, USA. The purpose of this study is to collect brain imaging and behavioral data from a transdiagnostic cohort of patients with common psychiatric diagnoses, as well as control participants.

Participants are recruited from the community via flyers, online advertisements and through patient referral from clinicals. All participants complete a clinical interview and an MRI scanning session. Participants were eligible for the study if they 1) were aged between of 18-35, 2) had no MRI contraindications, 3) were not colorblind, and 4) had no neurological abnormalities. All participants underwent Structured Clinical Interview for DSM-5 to determine psychiatric diagnosis. As a result, recruitment included both healthy individuals and individuals with a diverse set of clinical presentations, including affective and psychotic psychopathology.

### Additional Information on MRI processing and denoising

For the UK Biobank, we used the processed volumetric rs-fMRI data from the first imaging visit^81^. Each fMRI dataset was spatially normalised to MNI152 2-mm template space and FMRIB’s ICA-based X-noiseifier (FSL-FIX; ^82^) was trained on holdout set of participants and applied to the remaining participants to denoise the data. The mean global signal was extracted using a whole-brain mask and was regressed out of each dataset. A detailed outline of the processing, denoising and quality control of these data has been previously reported ^81^.

For the CNP data set, fmriprep v1.1.1^83^ was used. During this standardised and automated pipeline, each T1-weighted (T1w) volume was corrected for intensity non-uniformity using N4BiasFieldCorrection^84^ and skull-stripped using antsBrainExtraction.sh. Brain surfaces were reconstructed using recon-all from <u>FreeSurfer</u> v6. Spatial normalization to the MNI152 Nonlinear Asymmetrical template version 2009c was performed through nonlinear registration with ANTs^85^, using brain-extracted versions of both T1w volume and template. Brain-tissue segmentation of tissue classes was performed on the brain-extracted T1w using FSL FAST^86^. Functional MRI data were slice-time corrected using AFNI^87^ and realigned to a mean reference image using mcflirt^88^. Susceptibility distortion correction was performed by co-registering the functional image to the intensity-inverted T1w image with an representative EPI distortion atlas^89^. This was followed by co-registration to the corresponding T1w using boundary-based registration, implemented using FreeSurfer’s BBRegister. The motion-correcting transformations, field-distortion-correcting warp, BOLD-to-T1w transformation, and T1w-to-MNI warp were concatenated and applied in a single step using ANTS. ICA-based Automatic Removal Of Motion Artifacts (AROMA) was used to generate signal and noise and signal regressors for use in the non-aggressive variant of the method^90^. Regressors were calculated on the spatially smoothed output 6 mm FWHM kernel) and then applied to the unsmoothed pre-processed file. Following ICA-AROMA, we extracted mean time courses from eroded masks of the WM and CSF and regressed these signals out of the ICA-AROMA denoised data. Finally, each dataset was detrended with a 2nd order polynomial and high-pass filtered at 0.005 Hz using AFNI’s 3dTproject. The mean global signal was extracted using a whole-brain mask and was regressed out of each dataset. Further details on processing, denoising and quality control, please see are reported elsewhere^91^.

Both the HCP-EP and TCP datasets were acquired use the Human Connectome Project (HCP) MRI acquisition parameters. We therefore implemented the Minimal Processing Pipeline which was developed and optimized for HCP data^92^. The pipeline adapts steps from FMRIB Software Library (FSL; ^93^) and FreeSurfer to account for greater spatial and temporal resolution and HCP-data related distortions resulting from acquisition choices such as multiband acceleration, while aiming to remove the least amount of data necessary. During this pipeline, brain surfaces were reconstructed using recon-all from FreeSurfer v6. Skull stripped T1w and fMRI data were aligned using FSL Linear Image Registration Tool (FLIRT). Spin Echo EPI Field Maps with opposite phase encoding directions were used to estimate spatial distortion, using FSL topup and FLIRT was used to correct the scans for such distortions. This process was fine-tuned and optimised using FreeSurfer’s BBRegister. Functional MRI data realigned to a mean reference image using mcflirt^88^. Lastly, non-linear registration of Functional MRI data, aligned to individual’s structural volume space into standard MNI152 space was done using FLIRT and FMRIB’s nonlinear image registration tool (FNIRT). To denoise the fMRI data, ICA-FIX was implemented. During ICA-FIX, the fMRI data is decomposed into spatially independent components using Multivariate Exploratory Linear Optimized Decomposition into Independent Components (MELODIC). The resulting components are then classified as noise or signal. While ICA-AROMA uses a set of fixed rules depending on the time-course and frequency of each component, ICA-FIX uses a machine-learning based classifier. Here we used the pre-trained HCP_hp2000 classifier provided with ICA-FIX^82^, as the acquisition parameters of the fMRI data this classifier was train on are identical to those of the HCP-EP and TCP datasets. A temporal high-pass filter of 2000 was applied and a lenient threshold component labelling in FIX (t=10) was used. Finally, the mean global signal was extracted using a whole-brain mask and was regressed out of each dataset.

The steps described above resulted in processed and denoised fMRI dataset in MNI152 volume space for each individual. For each fMRI dataset, the time series were averaged within each of the 400 cortical^48^ and 19 subcortical^49^ parcels and pairwise Pearson’s correlations were computed to generate a 419□×□419 functional connectivity matrix, after which correlation values were z-scored and the upper-triangle of this matrix which consisted of 87,571 unique functional connectivity estimates were entered into the prediction models.

## References

1 Poldrack, R. A., Huckins, G. & Varoquaux, G. Establishment of best practices for evidence for prediction: a review. JAMA psychiatry 77, 534–540 (2020).

2 Varoquaux, G. Cross-validation failure: Small sample sizes lead to large error bars. Neuroimage 180, 68–77 (2018).

3 Whelan, R. & Garavan, H. When optimism hurts: inflated predictions in psychiatric neuroimaging. Biological psychiatry 75, 746–748 (2014).

4 Abramovitch, A., Short, T. & Schweiger, A. The C Factor: Cognitive dysfunction as a transdiagnostic dimension in psychopathology. Clinical Psychology Review 86, 102007 (2021).

5 East-Richard, C., R-Mercier, A., Nadeau, D. & Cellard, C. Transdiagnostic neurocognitive deficits in psychiatry: A review of meta-analyses. Canadian Psychology/Psychologie canadienne 61, 190 (2020).

6 McTeague, L. M., Goodkind, M. S. & Etkin, A. Transdiagnostic impairment of cognitive control in mental illness. Journal of psychiatric research 83, 37–46 (2016).

7 Millan, M. J. et al. Cognitive dysfunction in psychiatric disorders: characteristics, causes and the quest for improved therapy. Nature reviews Drug discovery 11, 141–168 (2012).

8 Shilyansky, C. et al. Effect of antidepressant treatment on cognitive impairments associated with depression: a randomised longitudinal study. The Lancet Psychiatry 3, 425–435 (2016).

9 Mohamed, S. et al. Relationship of cognition and psychopathology to functional impairment in schizophrenia. American Journal of Psychiatry 165, 978–987 (2008).

10 Green, M. F. Cognitive impairment and functional outcome in schizophrenia and bipolar disorder. Journal of Clinical Psychiatry 67, 3 (2006).

11 Diamond, A. Executive functions. Annual review of psychology 64, 135 (2013).

12 Fitzgerald, H. M. et al. Treatment Goals in Schizophrenia: A Real-World Survey of Patients, Psychiatrists, and Caregivers in the United States, with an Analysis of Current Treatment (Long-Acting Injectable vs Oral Antipsychotics) and Goal Selection. Neuropsychiatric Disease and Treatment 17, 3215 (2021).

13 McNaughton, E. C. et al. Patient attitudes toward and goals for MDD treatment: a survey study. Patient preference and adherence 13, 959 (2019).

14 Flechsig, P. in Die Localisation der geistigen Vorgänge insbesondere der Sinnesempfindungen des Menschen (De Gruyter, 1896).

15 Goldman-Rakic, P. S. Topography of cognition: parallel distributed networks in primate association cortex. Annual review of neuroscience 11, 137–156 (1988).

16 Margulies, D. S. et al. Situating the default-mode network along a principal gradient of macroscale cortical organization. Proceedings of the National Academy of Sciences 113, 12574–12579 (2016).

17 Voytek, B. et al. Oscillatory dynamics coordinating human frontal networks in support of goal maintenance. Nature neuroscience 18, 1318–1324 (2015).

18 Badre, D., Kayser, A. S. & D’Esposito, M. Frontal cortex and the discovery of abstract action rules. Neuron 66, 315–326 (2010).

19 Badre, D. & Nee, D. E. Frontal cortex and the hierarchical control of behavior. Trends in cognitive sciences 22, 170–188 (2018).

20 Elliott, M. L., Romer, A., Knodt, A. R. & Hariri, A. R. A Connectome-wide Functional Signature of Transdiagnostic Risk for Mental Illness. Biological Psychiatry 84, 452–459, doi:10.1016/j.biopsych.2018.03.012 (2018).

21 Romer, A. L. et al. Pervasively thinner neocortex as a transdiagnostic feature of general psychopathology. American Journal of Psychiatry 178, 174–182 (2021).

22 Lees, B. et al. Altered Neurocognitive Functional Connectivity and Activation Patterns Underlie Psychopathology in Preadolescence. Biological Psychiatry: Cognitive Neuroscience and Neuroimaging 6, 387–398, doi:10.1016/j.bpsc.2020.09.007 (2021).

23 Segal, A. et al. Regional, circuit, and network heterogeneity of brain abnormalities in psychiatric disorders. medRxiv (2022).

24 Cole, M. W., Repovš, G. & Anticevic, A. The frontoparietal control system: a central role in mental health. The Neuroscientist 20, 652–664 (2014).

25 Xia, C. H. et al. Linked dimensions of psychopathology and connectivity in functional brain networks. Nature Communications 9, 3003, doi:10.1038/s41467-018-05317-y (2018).

26 Vanes, L. D. & Dolan, R. J. Transdiagnostic neuroimaging markers of psychiatric risk: A narrative review. NeuroImage: Clinical 30, 102634 (2021).

27 Anticevic, A. et al. The role of default network deactivation in cognition and disease. Trends in cognitive sciences 16, 584–592 (2012).

28 Chopra, S. et al. Functional connectivity in antipsychotic-treated and antipsychotic-naive patients with first-episode psychosis and low risk of self-harm or aggression: a secondary analysis of a randomized clinical trial. JAMA psychiatry 78, 994–1004 (2021).

29 Vincent, J. L., Kahn, I., Snyder, A. Z., Raichle, M. E. & Buckner, R. L. Evidence for a frontoparietal control system revealed by intrinsic functional connectivity. Journal of neurophysiology 100, 3328–3342 (2008).

30 Huang, C.-C. et al. Transdiagnostic and illness-specific functional dysconnectivity across schizophrenia, bipolar disorder, and major depressive disorder. Biological Psychiatry: Cognitive Neuroscience and Neuroimaging 5, 542–553 (2020).

31 Fornito, A., Yoon, J., Zalesky, A., Bullmore, E. T. & Carter, C. S. General and specific functional connectivity disturbances in first-episode schizophrenia during cognitive control performance. Biological psychiatry 70, 64–72 (2011).

32 Baker, J. T. et al. Functional connectomics of affective and psychotic pathology. Proceedings of the National Academy of Sciences 116, 9050–9059, doi:10.1073/pnas.1820780116 (2019).

33 Dhamala, E., Jamison, K. W., Jaywant, A., Dennis, S. & Kuceyeski, A. Distinct functional and structural connections predict crystallised and fluid cognition in healthy adults. Human Brain Mapping 42, 3102–3118 (2021).

34 He, T. et al. Deep neural networks and kernel regression achieve comparable accuracies for functional connectivity prediction of behavior and demographics. NeuroImage 206, 116276 (2020).

35 Sripada, C. et al. Prediction of neurocognition in youth from resting state fMRI. Molecular psychiatry 25, 3413–3421 (2020).

36 Finn, E. S. et al. Functional connectome fingerprinting: identifying individuals using patterns of brain connectivity. Nature neuroscience 18, 1664–1671 (2015).

37 Jiang, R. et al. Gender differences in connectome-based predictions of individualized intelligence quotient and sub-domain scores. Cerebral cortex 30, 888–900 (2020).

38 Marek, S. et al. Reproducible brain-wide association studies require thousands of individuals. Nature 603, 654–660 (2022).

39 Schulz, M.-A. et al. Different scaling of linear models and deep learning in UKBiobank brain images versus machine-learning datasets. Nature communications 11, 1–15 (2020).

40 Helmer, M. et al. On stability of canonical correlation analysis and partial least squares with application to brain-behavior associations. BioRxiv, 2020.2008. 2025.265546 (2021).

41 Tong, X. et al. Transdiagnostic connectome signatures from resting-state fMRI predict individual-level intellectual capacity. Translational Psychiatry 12, 367, doi:10.1038/s41398-022-02134-2 (2022).

42 Boeke, E. A., Holmes, A. J. & Phelps, E. A. Toward robust anxiety biomarkers: a machine learning approach in a large-scale sample. Biological Psychiatry: Cognitive Neuroscience and Neuroimaging 5, 799–807 (2020).

43 Dhamala, E. et al. Proportional intracranial volume correction differentially biases behavioral predictions across neuroanatomical features and populations. bioRxiv (2022).

44 Smith, S. M. et al. A positive-negative mode of population covariation links brain connectivity, demographics and behavior. Nature neuroscience 18, 1565–1567 (2015).

45 Chen, J. et al. Shared and unique brain network features predict cognitive, personality, and mental health scores in the ABCD study. Nature communications 13, 1–17 (2022).

46 Miller, K. L. et al. Multimodal population brain imaging in the UK Biobank prospective epidemiological study. Nature neuroscience 19, 1523–1536 (2016).

47 He, T. et al. Meta-matching as a simple framework to translate phenotypic predictive models from big to small data. Nature Neuroscience, 1–10 (2022).

48 Schaefer, A. et al. Local-Global Parcellation of the Human Cerebral Cortex from Intrinsic Functional Connectivity MRI. Cerebral Cortex (New York, N.Y.: 1991) 28, 3095–3114, doi:10.1093/cercor/bhx179 (2018).

49 Fischl, B. et al. Whole brain segmentation: automated labeling of neuroanatomical structures in the human brain. Neuron 33, 341–355 (2002).

50 Ooi, L. Q. R. et al. Comparison of individualized behavioral predictions across anatomical, diffusion and functional connectivity MRI. NeuroImage 263, 119636 (2022).

51 Yeo, B. T. et al. The organization of the human cerebral cortex estimated by intrinsic functional connectivity. Journal of neurophysiology (2011).

52 Haufe, S. et al. On the interpretation of weight vectors of linear models in multivariate neuroimaging. Neuroimage 87, 96–110 (2014).

53 Tian, Y. & Zalesky, A. Machine learning prediction of cognition from functional connectivity: Are feature weights reliable? NeuroImage 245, 118648 (2021).

54 Chen, J. et al. There is no fundamental trade-off between prediction accuracy and feature importance reliability. bioRxiv (2022).

55 Váša, F. & Mišic, B. Null models in network neuroscience. Nature Reviews Neuroscience, 1–12 (2022).

56 Tozzi, L., Fleming, S. L., Taylor, Z. D., Raterink, C. D. & Williams, L. M. Test-retest reliability of the human functional connectome over consecutive days: identifying highly reliable portions and assessing the impact of methodological choices. Network Neuroscience 4, 925–945 (2020).

57 Zuo, X.-N. & Xing, X.-X. Test-retest reliabilities of resting-state FMRI measurements in human brain functional connectomics: a systems neuroscience perspective. Neuroscience & Biobehavioral Reviews 45, 100–118 (2014).

58 Zanto, T. P. & Gazzaley, A. Fronto-parietal network: flexible hub of cognitive control. Trends in cognitive sciences 17, 602–603 (2013).

59 Smallwood, J., Brown, K., Baird, B. & Schooler, J. W. Cooperation between the default mode network and the frontal–parietal network in the production of an internal train of thought. Brain research 1428, 60–70 (2012).

60 Allott, K. & Lin, A. in Risk Factors for Psychosis 269–287 (Elsevier, 2020).

61 Catalan, A. et al. Neurocognitive functioning in individuals at clinical high risk for psychosis: a systematic review and meta-analysis. JAMA psychiatry 78, 859–867 (2021).

62 Park, H.-J. & Friston, K. Structural and functional brain networks: from connections to cognition. Science 342, 1238411 (2013).

63 Petersen, S. E. & Sporns, O. Brain networks and cognitive architectures. Neuron 88, 207–219 (2015).

64 Bycroft, C. et al. The UK Biobank resource with deep phenotyping and genomic data. Nature 562, 203–209 (2018).

65 Lewandowski, K. E., Bouix, S., Ongur, D. & Shenton, M. E. Neuroprogression across the early course of psychosis. Journal of psychiatry and brain science 5 (2020).

66 Poldrack, R. A. et al. A phenome-wide examination of neural and cognitive function. Scientific data 3, 1–12 (2016).

67 Power, J. D. et al. Distinctions among real and apparent respiratory motions in human fMRI data. NeuroImage 201, 116041 (2019).

68 Fair, D. A. et al. Correction of respiratory artifacts in MRI head motion estimates. Neuroimage 208, 116400 (2020).

69 Power, J. D. et al. Methods to detect, characterize, and remove motion artifact in resting state fMRI. Neuroimage 84, 320–341 (2014).

70 Li, J. et al. Global signal regression strengthens association between resting-state functional connectivity and behavior. NeuroImage 196, 126–141 (2019).

71 Parkes, L., Fulcher, B., Yücel, M. & Fornito, A. An evaluation of the efficacy, reliability, and sensitivity of motion correction strategies for resting-state functional MRI. NeuroImage 171, 415–436, doi:10.1016/j.neuroimage.2017.12.073 (2018).

72 Weintraub, S. et al. Cognition assessment using the NIH Toolbox. Neurology 80, S54–S64 (2013).

73 Wechsler, D. Wechsler abbreviated scale of intelligence. (1999).

74 Singh, S. et al. The TestMyBrain digital neuropsychology toolkit: Development and psychometric characteristics. Journal of Clinical and Experimental Neuropsychology 43, 786–795 (2021).

75 Wechsler, D. Wechsler memory scale. (1945).

76 Wechsler, D. Wechsler adult intelligence scale. Archives of Clinical Neuropsychology (1955).

77 Raghunathan, T. E., Rosenthal, R. & Rubin, D. B. Comparing correlated but nonoverlapping correlations. Psychological Methods 1, 178 (1996).

78 Diedenhofen, B. & Musch, J. cocor: A comprehensive solution for the statistical comparison of correlations. PloS one 10, e0121945 (2015).

79 Dhamala, E., Yeo, B. T. & Holmes, A. J. Methodological Considerations for Brain-Based Predictive Modelling in Psychiatry. Biological Psychiatry (2022).

80 Chyzhyk, D., Varoquaux, G., Milham, M. & Thirion, B. How to remove or control confounds in predictive models, with applications to brain biomarkers. GigaScience 11 (2022).

81 Alfaro-Almagro, F. et al. Image processing and Quality Control for the first 10,000 brain imaging datasets from UK Biobank. Neuroimage 166, 400–424 (2018).

82 Salimi-Khorshidi, G. et al. Automatic denoising of functional MRI data: combining independent component analysis and hierarchical fusion of classifiers. Neuroimage 90, 449–468, doi:10.1016/j.neuroimage.2013.11.046 (2014).

83 Esteban, O. et al. fMRIPrep: a robust preprocessing pipeline for functional MRI. Nature Methods 16, 111–116, doi:10.1038/s41592-018-0235-4 (2019).

84 Tustison, N. J. et al. N4ITK: improved N3 bias correction. IEEE transactions on medical imaging 29, 1310–1320 (2010).

85 Avants, B. B., Tustison, N. & Song, G. Advanced normalization tools (ANTS). Insight j 2, 1–35 (2009).

86 Zhang, Y., Brady, M. & Smith, S. Segmentation of brain MR images through a hidden Markov random field model and the expectation-maximization algorithm. IEEE transactions on medical imaging 20, 45–57 (2001).

87 Cox, R. W. AFNI: software for analysis and visualization of functional magnetic resonance neuroimages. Computers and Biomedical research 29, 162–173 (1996).

88 Jenkinson, M., Bannister, P., Brady, M. & Smith, S. Improved optimization for the robust and accurate linear registration and motion correction of brain images. Neuroimage 17, 825–841 (2002).

89 Wang, S. et al. Evaluation of field map and nonlinear registration methods for correction of susceptibility artifacts in diffusion MRI. Frontiers in neuroinformatics 11, 17 (2017).

90 Pruim, R. H. R. et al. ICA-AROMA: A robust ICA-based strategy for removing motion artifacts from fMRI data. NeuroImage 112, 267–277, doi:10.1016/j.neuroimage.2015.02.064 (2015).

91 Aquino, K. M., Fulcher, B. D., Parkes, L., Sabaroedin, K. & Fornito, A. Identifying and removing widespread signal deflections from fMRI data: Rethinking the global signal regression problem. Neuroimage 212, 116614 (2020).

92 Glasser, M. F. et al. The minimal preprocessing pipelines for the Human Connectome Project. Neuroimage 80, 105–124 (2013).

93 Jenkinson, M., Beckmann, C. F., Behrens, T. E., Woolrich, M. W. & Smith, S. M. FSL. Neuroimage 62, 782–790, doi:10.1016/j.neuroimage.2011.09.015 (2012).

